# Regional Prevalence, Stage Distribution, and Temporal Trends of Cardiovascular-Kidney-Metabolic Syndrome in the Americas, Europe and Western Pacific: A Systematic Review and Meta-Analysis

**DOI:** 10.1101/2025.06.04.25328945

**Authors:** Jiajin Li, Lei Lu, Yizhu Zhang, Qingze Gu, Tingting Zhu, Fengshi Jing, Karen Tu, Peter Tanuseputro, Dijing You, Hao Ren, Weibing Cheng, Qingpeng Zhang, Jiandong Zhou

**Author notes:** Correspondence to: Jiandong Zhou, PhD, Department of Family Medicine and Primary Care, Li Ka Shing Faculty of Medicine, The University of Hong Kong, Hong Kong SAR, China; School of Public Health, Li Ka Shing Faculty of Medicine, The University of Hong Kong, Hong Kong SAR, China; Department of Pharmacology and Pharmacy, The University of Hong Kong, Hong Kong SAR, China.

## Abstract

**Background:** The cardiovascular-kidney-metabolic (CKM) syndrome, first proposed by the American Heart Association (AHA) in 2023, represents a groundbreaking conceptual framework that integrates these three interrelated conditions into a unified clinical entity. Despite growing research on its prevalence, risk factors, and clinical management, the regional burden of CKM syndrome remains poorly characterized. To address this gap, we aimed to estimate the prevalence of CKM syndrome, providing critical insights into the regional impact of this novel disease definition.

**Methods:** In this study, we conducted literature retrieval in both English (PubMed, Web of Science and Wiley Online Library) and Chinese databases (CNKI and Wangfang), as well as the journal official websites (e.g. American Heart Association (AHA) and American Society of Nephrology (ASN)) from database inception until January 20, 2025, followed by an update search until April 1, 2025. Grey literature such as posters and preprint articles, and citations from the identified reviews were also searched for. Cross sectional and cohort studies were included without language limitation. Studies employing other study designs or were done in people who were not representative of the general population (e.g. people with specific diseases) were excluded. Summary data were obtained from included studies. The primary outcomes were the prevalence of CKM syndrome and its different stages (stages 0 - 4) among general population. The combined prevalence was obtained with Freeman-Tukey Double Arcsine Transformation method. The estimated annuls percentage change (EAPC) was employed to explore the trend of CKM syndrome. This study is registered with PROSPERO (CRD420251037912).

**Findings:** From 2,708 identified 2,708 related articles, 28 studies with 29 datapoints, encompassing 1,561,209 individuals, were included. The overall pooled prevalence of CKM syndrome (Stages 1–4) in the general population was 0.88 [95% CI 0.86–0.91]. This estimate was 0.85 [95% CI 0.76–0.91] in a sensitivity analysis selecting one representative study per database to test the magnitude of potential duplicate bias. The combined prevalence of stages 1, 2, 3 and 4 was 0.23 [95% CI 0.19–0.27], 0.46 [95% CI 0.41–0.51], 0.08 [95% CI 0.05–0.11], 0.07 [95% CI 0.04–0.12], respectively, displaying as the Stage 2 patients were the majority of CKM syndrome. The EAPC of CKM syndrome in the period of 1991-2021 was (-0.55% [95% CI -0.90 to 0.21], p=0.0024), displaying a significant decreased trend. Stratified by countries, the pooled estimates were 0.91 [95% CI 0.90–0.93] for USA, 0.90 [95% CI 0.87–0.93] for China, and 0.77 [95% CI 0.69–0.84] for other countries (UK, Italy and South Korea). CKM syndrome prevalence demonstrated an increasing trend with a higher proportion of males (male/female ratio <0.98) and with increasing mean age (up to 56.5 years). Statistically significant disparities were observed across social development index (SDI) level, data source and countries.

**Interpretation:** This study provides the pooled regional prevalence of CKM syndrome in the general population; these findings are valuable for understanding the current burden of CKM syndrome and facilitating more research into the clinical management and prevention. While a slight decreasing temporal trend was observed based on the included studies, the relatively high combined prevalence suggests more epidemiological research into missing regions, such as Africa and South America, to verify this finding.

**Research In Context:** *Evidence before this study:* Cardiovascular-Kidney-Metabolic (CKM) syndrome has drawn substantial attention to the complex interactions among cardiovascular, kidney and metabolic diseases. Previous studies reported a heavy burden of CKM syndrome in certain jurisdictions, for example, the weighted prevalence was 83.7% in China in 2019 and 79.5% in the USA in 2023 nationwide. However, the epidemiological studies reporting the burden of CKM syndrome regionally remain inadequate. Thus, we did a meta-analysis to map the prevalence of CKM syndrome globally. We searched PubMed from database inception to April 1, 2025, using terms (with variants) like “meta-analysis”, “CKM syndrome” and “prevalence”, and did not find any other meta-analysis reporting this topic.

*Added value of this study:* This study provides a comprehensive regional view of the prevalence of CKM syndrome in the general population, with additional temporal trend analysis based on study period, male/female ratio and average age. Our analysis indicated a significant but slight downward trend of the prevalence of CKM syndrome from 1991 to 2021. The pooled prevalence of CKM syndrome was between 0.85 [95% CI 0.76–0.91] and 0.88 [95% CI 0.86–0.91], with stage 2 patients taking the highest proportion. Significant differences were observed across sociodemographic index (SDI), data source and countries. Middle level SDI, population-based design and USA showed the highest prevalence of CKM syndrome compared to their counterparts.

*Implications of all the available evidence:* This study provides up-to-date estimates of regional burden of CKM syndrome and can be referred to future public health promotion planning, as well as to explore the risk factors and locate the high-risk population. The relatively high pooled prevalence of CKM syndrome, nearly close to 90%, highlights the urgent need for developing risk stratification and prevention strategies for CKM syndrome patients, especially preventing staging progress among substantial stage 2 patients. The significant disparities in the prevalence of CKM syndrome across SDI, data source of the study and countries necessitating targeted strategies and further study design of this topic. Both developed and developing countries are facing heavy burden of CKM syndrome, where public health policies to reduce risk factors of CKM syndrome might be effective in alleviating the burden. Although the reported prevalence of CKM syndrome has declined from 1991 to 2021, this trend may be influenced by publication bias, as key regions with potentially higher CKM syndrome risks, such as Africa and South America, lack sufficient prevalence data. Therefore, more studies need to be conducted in these regions for more accurate estimate of global prevalence. Besides, more population-specific prevalence (such as sex, age group, different groups of patients) are also needed to help with risk factors determination.

## Introduction

In the wave of population aging, cardiovascular diseases (CVDs) have become a major global public health problem. Aging is related to gradual and persistent inflammation as well as oxidative stress in cardiovascular system, which ultimately increases the susceptibility of CVDs.^1^ By age 70, over 70% of adults are estimated to develop CVDs, often accompanied by multimorbidity such as chronic kidney disease (CKD) and metabolic disorders like diabetes and gout.^2,3^ The incidence of multiple chronic conditions increases the risk of all-cause mortality, with CVDs being one of the greatest impact factors.^4^

To better describe the complex interaction of cardiovascular, chronic renal and metabolic disorders, the American Heart Association recently introduced a new clinical construct: “Cardiovascular Kidney Metabolic (CKM) Syndrome” ^5^. The CKM Syndrome is defined as a progressive condition characterized by the interrelated dysfunction of the cardiovascular system, kidneys, and metabolic processes, driven by shared risk factors including inflammation, oxidative stress, and lifestyle-related exposures. This concept describes a spectrum of pathophysiological interactions among CVDs, CKD, and metabolic abnormalities, ultimately leading to multiorgan dysfunction and challenging treatment complexity.^5^ CKM is staged from 0 to 4 based on clinical burden, with Stage 0 indicating no identifiable risk where maintaining healthy lifestyle is essential for preventing disease progression, Stage 1 standing for excess adiposity, Stage 2 indicating metabolic risk factors, Stage 3 mainly focusing on subclinical CKD in CKM syndrome patients and Stage 4 representing overt cardiorenal disease with established complications.^6^

Recent studies showed a considerable burden of this syndrome in some countries. Recent studies showed considerable burden of this syndrome in some countries. In the USA, almost 90% of adults meet the criteria of CKM syndrome (stage 1 or higher) with 15% in advanced stages (stage 3 or 4), yet little improvement has been observed from 2011 to 2020. Similarly, in China, only 10.5% of people aged over 45 were in good CKM health (stage 0), while 27.2% experience CKM deterioration within four years.^7^ Despite lacking of global estimate of CKM syndrome burden, these findings underscore the pressing global health challenges posed by CKM syndrome and its implications in severe aging regions like Hong Kong, Japan and South Korea.

CKM syndrome can develop when effectively treated CVDs are followed by the emergence of renal and metabolic comorbidities over decades of survival. Notably, individuals with CVDs also face an elevated cancer risk compared to non-CVD populations, particularly those with atherosclerotic CVDs (Hazard Ratio 1.20). Shared risk factors, including chronic inflammation and smoking, further link CVDs, CKD, metabolic disorders, and cancer.^8-10^ Considering the significant impact of CKM syndrome on individual health, numerous epidemiological studies have been conducted to explore its prevalence.^11-13^

However, these studies mostly have only focused on a particular location or population, such as diabetes patients and healthcare workers, rather than exploring the temporal, gender, age patterns of CKM syndrome prevalence in a regional scale. Recently, a study using the Global Burden of Disease (GBD) 2021 platform explored the burden caused by CKM syndrome from 1990 to 2021, indicating that socioeconomic development, age, location and time are key influencing factors of the related burdens.

Given the substantial body of published epidemiological research on CKM syndrome since 2023, a meta-analysis based on real-world studies that examine the regional prevalence of CKM syndrome is warranted to promote the development of preventive and clinical management strategies. Thus, the main goal of this systematic review and meta-analysis was to investigate the prevalence and temporal trends of CKM syndrome in major countries, as well as the variation based on age, sex, study period, sociodemographic status and basic study settings.

## Methods

### Search Strategy and Selection Criteria

In this literature review and meta-analysis, two authors (J.J. Li and J.D. Zhou) independently searched for relevant studies in English databases (PubMed, Web of Science and Wiley Online Library, and Google Scholar), Chinese databases (CNKI and Wanfang), and directly from the official websites of journals (e.g. ASN and AHA). The search spanned from the inception of each database to January 20, 2025, using Medical Subject Headings (MeSH) and free-text terms related to CKM syndrome and its prevalence. An updated search was conducted until April 1, 2025. No language restrictions were applied; non-English articles were translated with manual verification to ensure accuracy. Detailed search strategies are provided in **(Table S1; supplementary pp 1-2)**. In addition, we checked the citations from the identified reviews during the literature searching. We also searched for grey literature though Google search engine.

Studies were included in this meta-analysis if they: (1) employed a cross-sectional or cohort design; (2) recruited participants from the general population or community settings; (3) provided sufficient data to calculate the prevalence of CKM syndrome with its 95% confidence interval (CI). Studies were excluded if they: (1) used selected samples (such as patients with diabetes or hospital-based cohorts); (2) had a sample size of fewer than 100 participants, given the potential for bias from small samples;^14^ (3) were review articles, case reports, commentaries, or reports; and (4) were solely qualitative. During our literature review, we identified a group of related articles utilizing the same public databases (e.g., UK Biobank and NHANES). Traditionally, to avoid duplication bias, only the most representative article from such a group is selected. However, a recent article redefined the concept of duplicate articles and proposed a decision tree for identifying duplicates in the context of widely used public databases.^15^ Applying this framework, we determined that none of the articles using the same databases were duplicates, leading us to include all of them in our analysis. Additionally, given the novelty of this framework, we decided to compare the combined prevalence of CKM syndrome across all included articles with the prevalence reported by a single representative article for each database. For this purpose, we defined a "representative article" as one with the largest sample size, comprehensive prevalence data across all stages of CKM syndrome, the longest time frame, and a high-quality assessment. This comparison aims to preliminarily evaluate the reliability of the new decision framework. Also, we further use the latter one to explore the pooled prevalence of each CKM syndrome stage to avoid potential duplicate publication bias.^15^ Moreover, studies reporting prevalence for two or more distinct cohorts within a single article were treated as separate studies in the meta-analysis.

All included articles were imported into EndNote (version 21.5) for reference management, and duplicates were removed. Discrepancies between the two reviewers were resolved by involving a third author (LL) to reach consensus. When essential data were missing from an article, we contacted the corresponding author to obtain the necessary information. The study was registered with PROSPERO (registration number: CRD420251037912).

### Data analysis

Data extracted from the included studies comprised the following: (1) first author and publication year; (2) study location, including countries, WHO regions, Socio-demographic Index (SDI), and Human Development Index (HDI); (3) study time interval, data source, study design, and staging criteria; (4) participant characteristics, such as male/female ratio (M/F ratio) and average age; and (5) total prevalence of CKM syndrome, prevalence by stage, their 95% CIs, and sample size. Detailed information on SDI and HDI, both comprehensive indicators of a country’s socioeconomic development, is provided in **(Text S1; supplementary pp 10)**. When prevalence estimates were not directly reported, they were calculated from available data within the articles. Due to the scarcity of sex- and age-specific prevalence, we used the ratio between the number of males and females (M/F ratio) and average age in the study as the proxy of sex and aging, to primarily explore their roles in the burden of CKM syndrome **(Text S2; supplementary pp 10)**. Studies employing different staging descriptions were classified into three categories: Criteria 1, Criteria 2 and Criteria Not Stated **(Text S3; supplementary pp 11)**.

We evaluated study quality using the Joanna Briggs Institute Critical Appraisal Checklist for Prevalence Studies.^16^ The overall quality of this meta-analysis was assessed according to the Grading of Recommendations Assessment, Development, and Evaluation (GRADE) framework.^17^.^17^

The pooled prevalence of CKM syndrome (total of stage 1 to 4 and by stages) and its 95% CI were obtained using the random effect models with Freeman-Tukey Double Arcsine Transformation, which was used for stabilizing the variances and providing more accurate combined results in the case of rates close to margins existing.^18^ Heterogeneity was estimated using the Q and I^2^ statistics, and I^2^ larger than 50% means substantial heterogeneity. Potential sources of heterogeneity were explored with subgroup analysis and meta-regression. Subgroup analysis was based on following variables: WHO regions, HDI levels, SDI levels, databases, study period, sex ratio, sample size, study design, staging criteria and data source. Subgroup analysis was based on studies that provided all-age prevalence, across sexes. To maintain comparable number of studies within the subgroup analysis, study period was divided into six intervals: 1991-2003, 2004-2007, 2008-2011, 2012-2015, 2016-2018 and 2019-2021. Subgroup differences were assessed using the χ^2^ test. Meta-regression was employed to further explore the influence of age, sex ratio and sample size. We performed sensitivity analyses to evaluate the robustness of results by computing how the overall effect size would change by removing one study at a time or removing studies with low quality. Publication bias was assessed by funnel plots and the Egger’s test for funnel plot asymmetry.

Estimated annual percentage change (EAPC) was calculated to explore the temporal trends of CKM syndrome prevalence, as well as trends based on the male/female ratios and mean values of age of participants **(Text S4; supplementary pp 11)**. Locally weighted linear regression (LOESS) was used to generate a moving average for scatterplot smoothing among the datapoints. If lacking of significance in the full-scale analysis, we would look for thresholds of male/female ratio and average age of participants that distinguishes between statistically significant and non-significant differences using exhaustive enumeration approach, to uncover potential hidden associations that might be masked when considering the entire datasets. And the results can be referred to future study design, such as constructing cohorts with similar population tributes for more efficiently getting meaningful findings.

Our analysis was performed with R (version 4.2.2) and the included meta-analytic models were fitted using the R package meta. Statistical significance was set at a two-sided p value of less than 0·05. We followed PRISMA reporting guidelines.^19^ **(Table S4; supplementary pp 6-9)**

## Results

Of 2,708 records identified, 2,223 articles were inspected for title, abstract and 48 articles were gone through full-text screening to determine eligibility. Finally, 29 datapoints of 28 articles from 6 major jurisdictions were found to be eligible for further analysis **(Figure 1)**.^20-47^ The main exclusion reasons include insufficient data, limited research population and selected samples. All of the included studies were reported in English and details can be referred to **Table 1** and **Table 2**. Almost all of the studies encompassed each stage’s prevalence, except 8 studies that only reported the first three stages’ prevalence or combined the prevalence of stage 3 and stage 4.^26,27,32,35,36,38,45,46^ Only 5 articles reported both all-age and age-specific prevalence for CKM syndrome, but the age intervals varied.^20,37,43,45,47^ All included studies involved 1,561,209 participants and 1,251,524 CKM patients from 6 major jurisdictions in 3 WHO regions. Those reported age-specific prevalence encompassed 171,605 participants and 147,443 patients from 3 major countries in Americas and Western Pacific. In addition, 11 studies reported sex-specific prevalence,^20,21,23,24,26,29,31,37,43,45,47^ 5 studies explored ethnicity-based prevalence,^20,21,24,37,47^ and 7 studies reported temporary trend of CKM prevalence.^20,23,24,26,29,37,43^ Furthermore, three studies reported detailed prevalence considering other factors such as weight status, household income and smoking status.^43,45,47^ The results of quality assessment of included articles are provided in **(Table S2; supplementary pp 3)**.

**Table 1.**
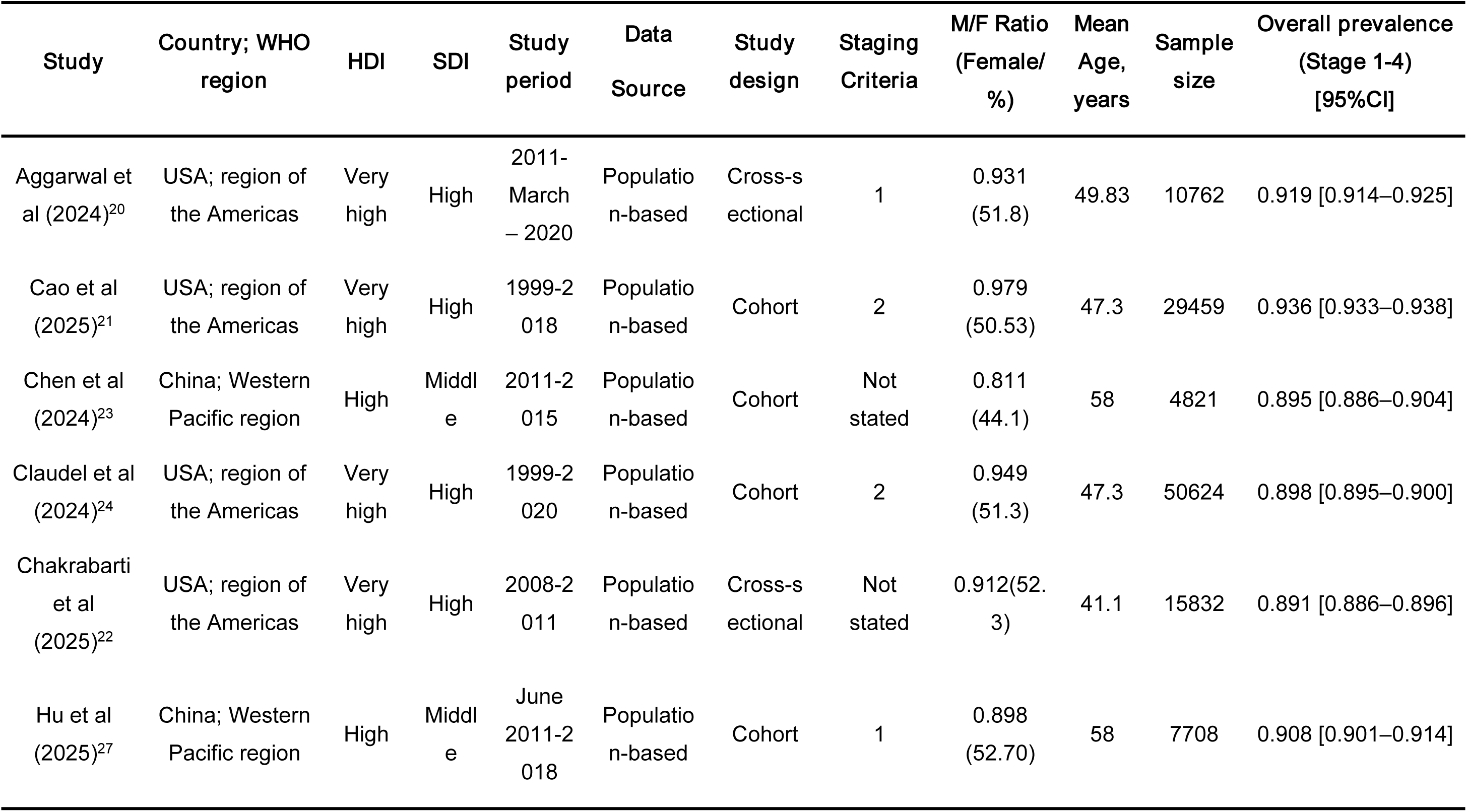
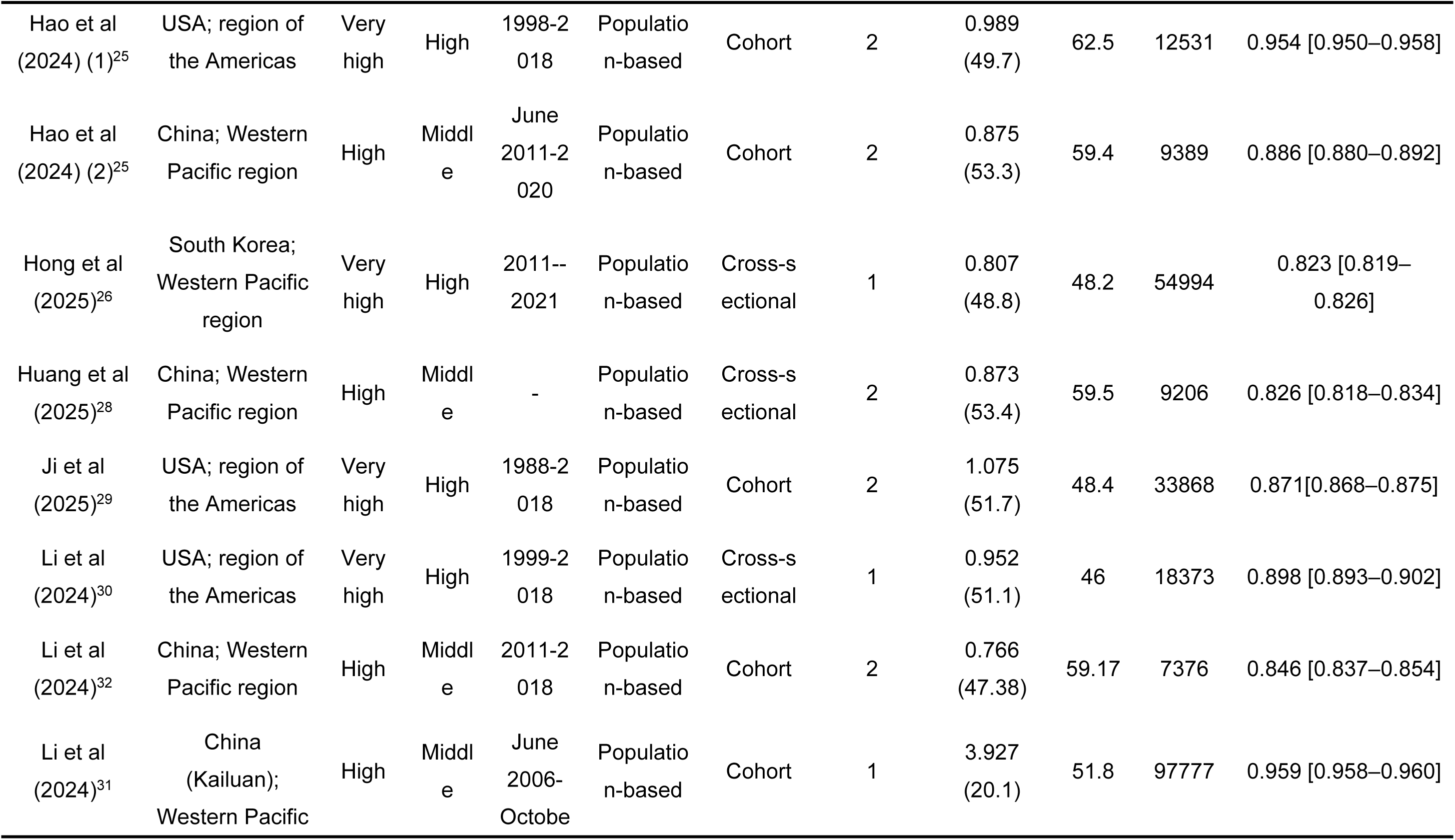
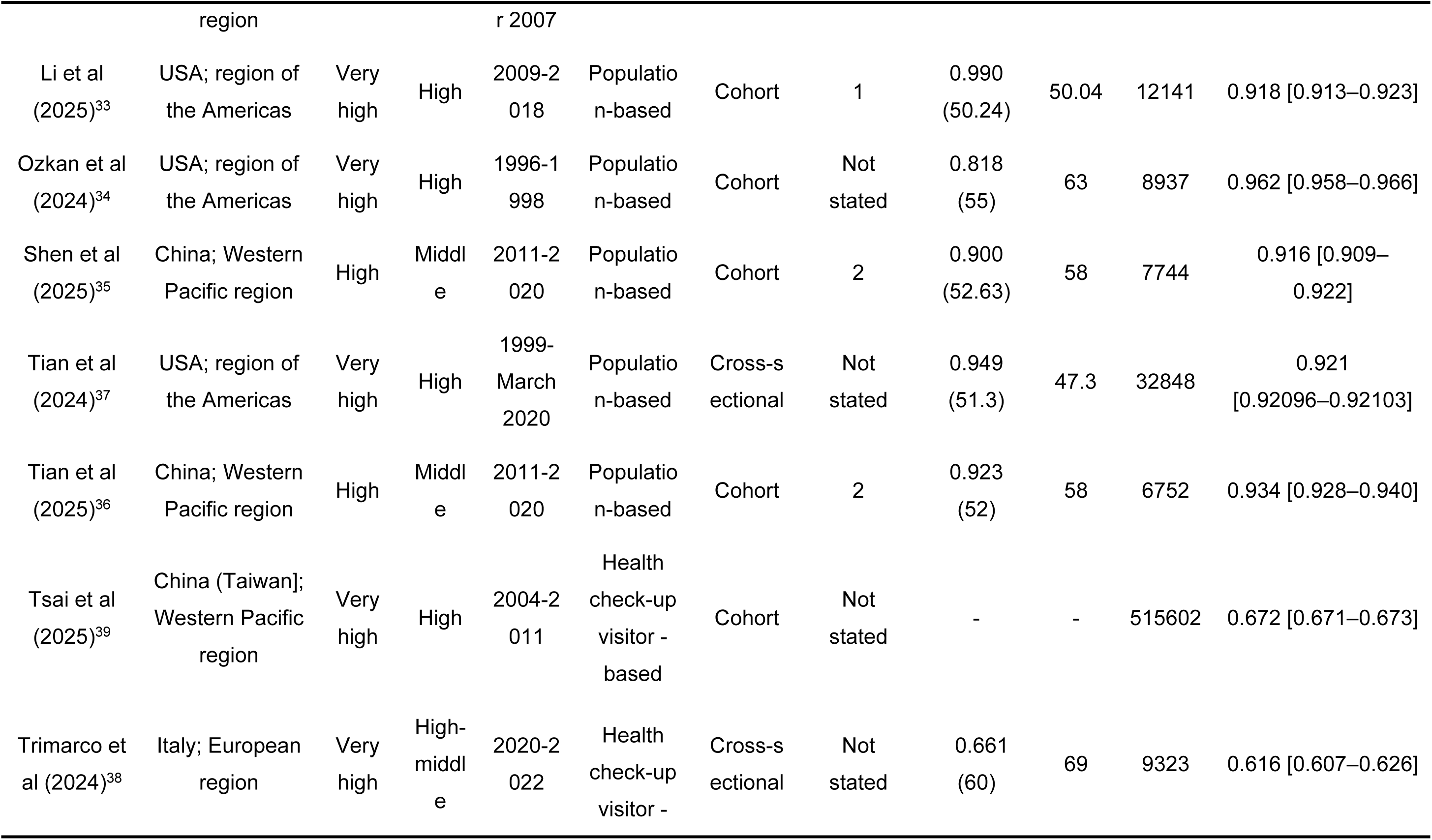
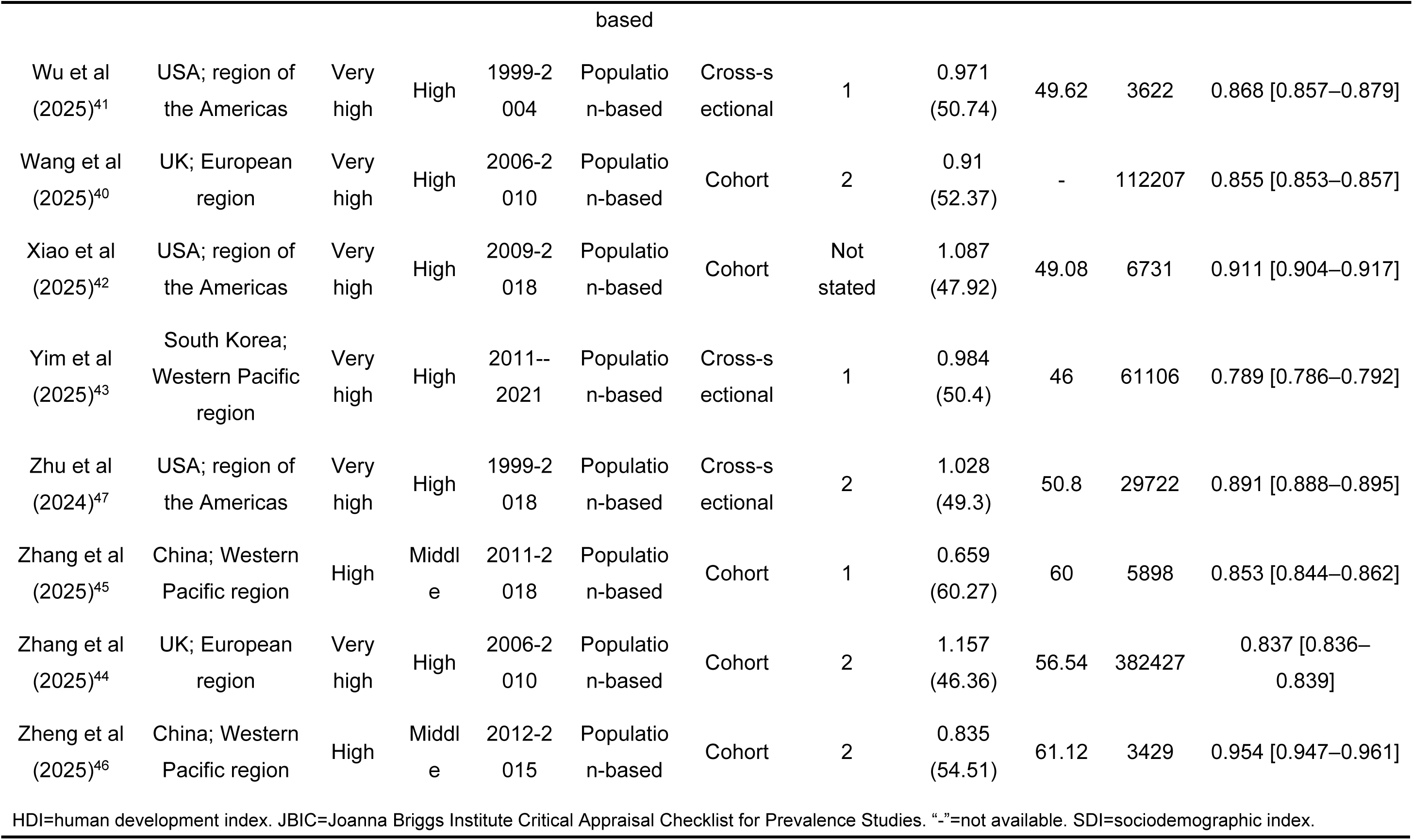
Basic information and quality of included articles.

**Table 2.**
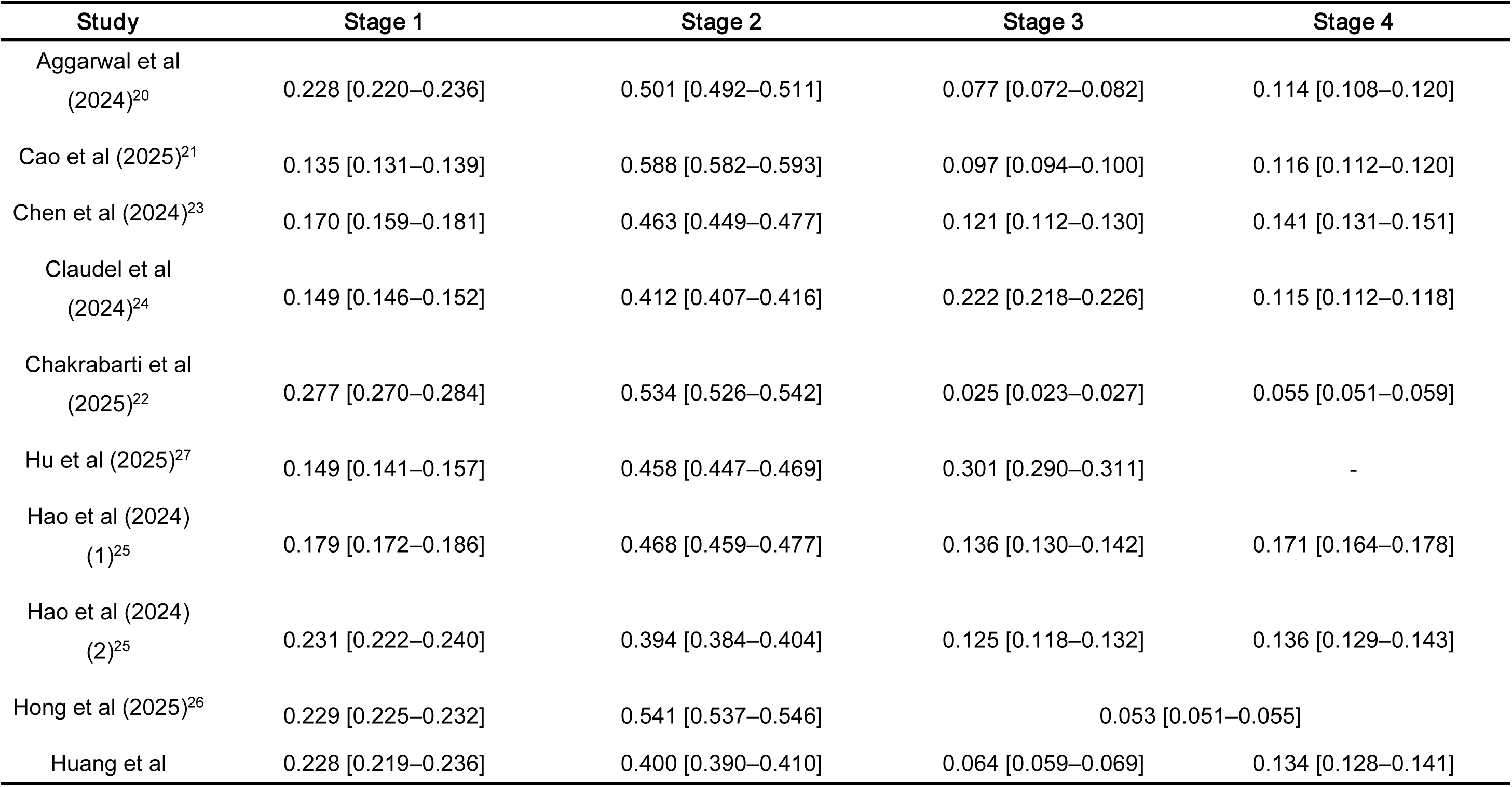
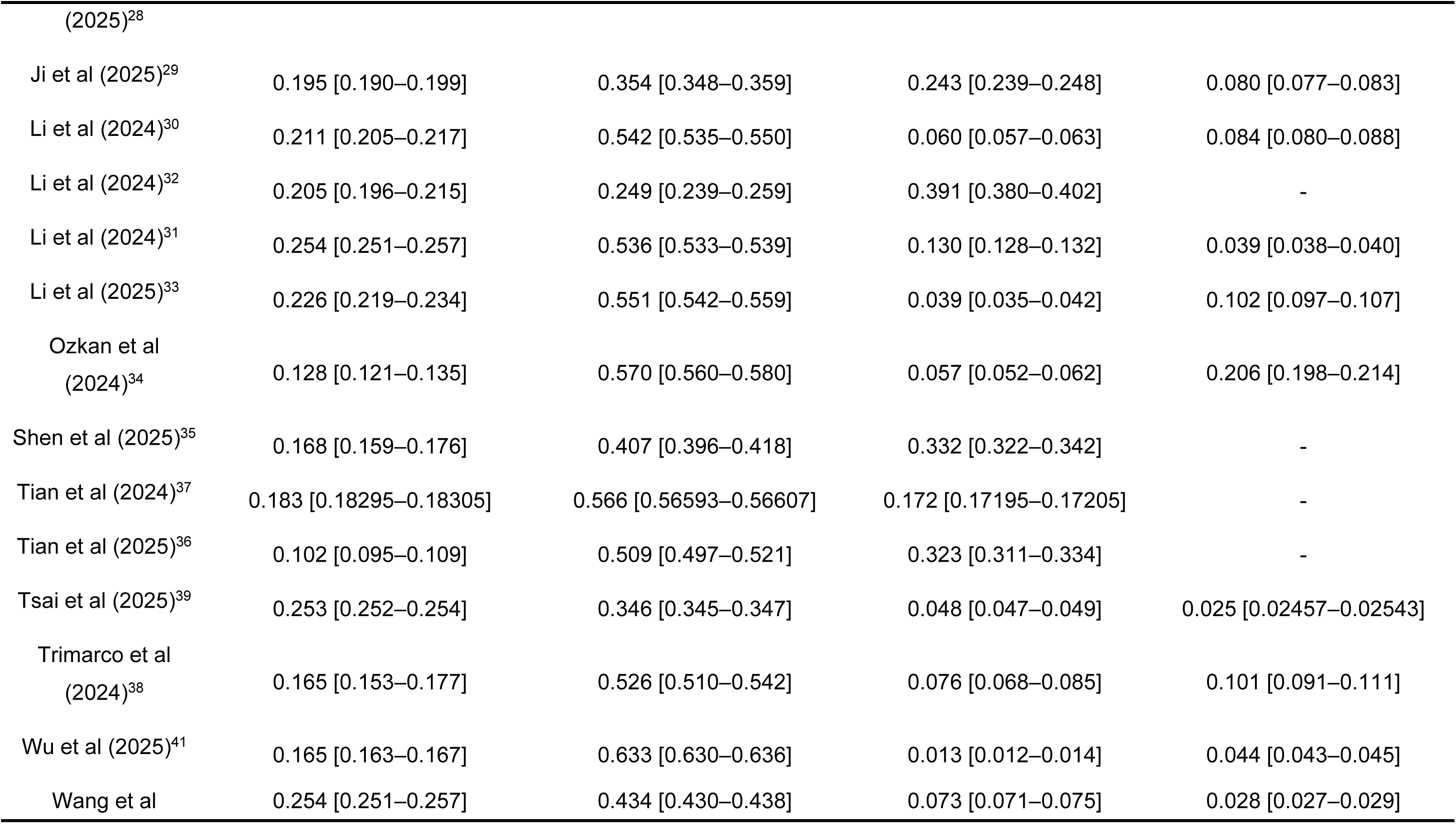
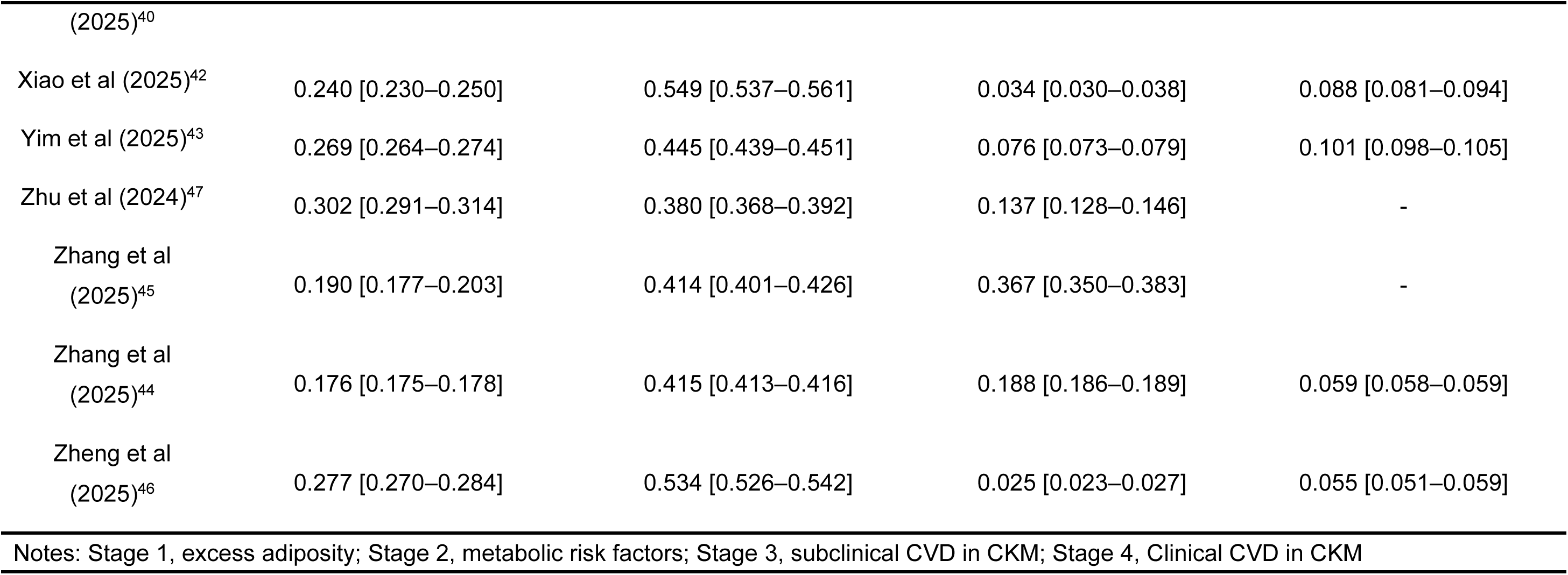
CKM syndrome prevalence by stages of included studies [Prevalence (95% CI)]

The pooled prevalence of CKM syndrome (Stages 1 to 4) was 0.88 [95% CI 0.86–0.91] across three WHO regions **(Figure 2)**. The National Health and Nutrition Examination Survey (NHANES)^20,21,24,25,29,30,33,37,41,42,47^ and China Health and Retirement Longitudinal Study (CHARLS)^23,25,27,28,32,35,36,45,46^ were two most used public databases regarding to CKM syndrome epidemiological research. After selecting one highest quality articles from each database the combined prevalence dropped to 0.85 [95% CI 0.76–0.91]. Stratified by stages, the pooled prevalence is 0.23 [95% CI 0.19–0.27], 0.46 [95% CI 0.41–0.51], 0.08 [95% CI 0.05–0.11] and 0.07 [95% CI 0.04–0.12], respectively **(Figure 3)**.

From 1991 to 2021, the prevalence of CKM syndrome exhibited a decreasing trend with survey periods (β = -0.006 [95% CI -0.009 to -0.002], p = 0.002) and its corresponding EAPC was -0.55% [95% CI -0.90 to -0.21]. Specifically, the EAPCs of CKM syndrome were 4.11% [95% CI -2.57–11.24] and -0.03% [95% CI -0.56–0.49] for different male/female ratios and average age of participants, respectively, but both were not statistically significant. **(Figure 4)**. When including only data points with a male/female ratio below 0.98 in the analysis, the prevalence of CKM syndrome exhibited a statistically significant increasing trend (β = 0.614 [95% CI 0.212–1.015], p = 0.005), with an estimated annual percentage change (EAPC) of 84.72% [95% CI 23.67–175.89]. Similarly, when restricting the analysis to included articles with a mean age younger than 56.5 years, the prevalence also showed a statistically significant increasing trend (β = 0.013 [95% CI 0.001–0.025], p = 0.030), with an EAPC of 1.31% [95% CI 0.14–2.50].

The subgroup analysis refers to **Figure 5**. Notably, significance disparity was observed in the combined prevalence of CKM syndrome stratified by SDI levels (p = 0.02), where areas with middle SDI level had slightly higher prevalence than those with high SDI level. In terms of data source, the pooled prevalence significantly varied (p < 0.01), with population-based studies considerably higher than those with health check-up visitors-based (clinic-based). Additionally, considerable differences (p < 0.01) occurred in different countries, with USA displaying the highest combined prevalence 0.91 [95% CI 0.90–0.93], followed by China 0.90 [95% CI 0.87–0.93]. Furthermore, though no statistical significance was observed among different study periods, we can still find a decreased pattern of pooled prevalence, with 1991-2003 0.92 [95% CI 0.89–0.95] showed the highest prevalence of CKM syndrome, while 2019-2021 0.80 [95% CI 0.71–0.88] showed the lowest one, which is consistent with the above-presented temporal trend analysis results.

In terms of sensitivity analysis, the pooled prevalence was not greatly affected after exclusion of low-quality studies **(Figure 2)**. The leave-one-out sensitivity analysis captured the study by Tsai that was influential for the combined prevalence **(Figure S1; supplementary pp 12)**. Funnel plots display substantial publication bias in the combined prevalence of CKM syndrome, which was consistent with the result of Egger’s test **(p = 0.003; Figure S2; supplementary pp 13)**. The certainty of evidence with GRADE approach was concluded as low for the prevalence of CKM syndrome **(Table S3; supplementary pp 5)**.

## Discussion

Our analysis suggested a slight downward trend in CKM syndrome prevalence between 1991 and 2021, which decreased significantly in the period of before 2019. The polled all-age prevalence was 0.88 [95% CI 0.86–0.91] with all included studies and dropped to 0.85 [95% CI 0.76–0.91] after selecting representative articles for each database. Significant disparities in the prevalence of CKM syndrome were observed among China, USA and other major countries. Moreover, significant upward trends were observed as the increase of sex ratio till 0.98 and average age till 56.5 years. Countries with high or high-middle SDI displayed a relatively high prevalence of CKM syndrome, though the highest prevalence of CKM syndrome was observed in middle SDI regions.

Even though there are no other studies estimating the regional prevalence of CKM syndrome, the results found in this study was similar to the estimating of several key components (such as high BMI and diabetes) of CKM syndrome using GBD 2021.Both of the studies displayed substantial burden of CKM syndrome, and Western Pacific region (such as China) facing the largest burden. However, the estimated temporal trend is inconsistent with the upward trend attained in the study of GBD 2021. We suggested several reasons for this inconsistency. Firstly, the data in Africa, South America and South Asia was missing in this study, where higher risks of CKM syndrome may exist.^48^ Thus, the estimation in this study might be lower than actual. Secondly, this disease was only proposed in 2023, and the indicators in its classification criteria across several aspects, for example, basic health indicators (BMI, waist circumference, blood pressure), metabolic syndrome (diabetes and hypolipidemia), CVD risk prediction equation (which also requires some socioeconomic information of the participants), CKD risk prediction and the medical history of CVD and CKD.^5^. Besides, most of the existing studies utilize databases from 2021 or even earlier. As a result, when including research subjects, a certain portion of the population will be excluded due to the absence of a specific indicator. This will lead to a certain degree of selection bias, thereby underestimating the prevalence of CKM syndrome due to potential loss of a portion of CKM syndrome patients.^49^

Despite the trend analysis, the global prevalence of CKM syndrome remains relatively high, among which stage 2 is the majority. This is also consistent with the finding in the study of GBD 2021, which high BMI and diabetes (both are in the stage 2 classification criteria) were the major contributors to years lived with disability (YLD) due to CKM syndrome and projected continuously increased through 2046. The classification of stage 2 mainly involves metabolic syndrome. The rising of metabolic syndrome has been observed in the early 2000s and it is mainly driven by unhealthy lifestyle, such as poor sleep quality and lack of physical activities. Latest investigations in major countries such as China and USA all reported the prevalence of metabolic syndrome of around 30–35% nationwide. As the medical care for metabolic syndrome has been developed for a long time, it improves the survival for metabolic syndrome patients, which eventually constitute the major part of CKM syndrome patients. Public health approaches targeted at metabolic syndrome patients might be effective in mitigating the burden of CKM syndrome.

Significant disparity was observed across different SDI levels, with the middle level of SDI displayed the highest prevalence of CKM syndrome. The result is logical because it is known that better health is positively linked with socioeconomic status. Even though related risk factors such as environmental pollutions, dietary habits, aging are increasing with the socioeconomic development, developed countries may be more experienced in regulating these adverse effects than developing countries, eventually displaying lower burden of CKM syndrome.^50^ However, this result might be biased due to possibly under-reporting of CKM syndrome cases in countries with lower level of SDI.^14^ Besides, prevalence estimates obtained using population-based and health check-up visitor-based (clinic-based) approaches were significantly different, with the former one displaying the highest prevalence. This may suggest potential selection bias from the latter method, because amount of people possibly did not participate in the program due to personal reasons or obstacles brought by the CKM syndrome itself, such as impaired mobility or hospitalization of the latter stage patients.

The upward trend of CKM syndrome with the increase of sex ratio (male versus female) below 0.98, is potentially attributed to the complex interaction among sex hormones and several key components of CKM syndrome.^29^ For example, estrogen displays a protective role against kidney injury while testosterone lies in the pathogenesis of chronic kidney diseases.^51^ Besides, it was observed that the increment of Stage 3 prevalence is more pronounced in males rather than females, and it was explained by the fact that males accumulate adiposity more easily than females.^52^ Fat accumulation is associated with increased risk of diabetes and chronic kidney disease, which belong to the major component of CKM syndrome. The prevalence of CKM syndrome also showed upward trend along with the average age of participants, but also limited in the range below 56.5 years. CKM syndrome belongs to comorbidity, thus the result is consistent with most of the studies about aging and comorbidity, where aging is a major risk factor.^53^ In this study, there are no simple linear relationships between age, sex and prevalence of CKM syndrome, suggesting that more studies about sex difference and ages are needed to deepen the understanding of the basic biological features in the progression of CKM syndrome.

Nonetheless, this study has some limitations. Firstly, epidemiological investigations of CKM syndrome in some jurisdictions were missing, particular in low SDI countries, which might influence the representative of the estimate in this study. Compared to other large scale meta-analysis about prevalence estimation, data source was limited in this study, especially the lack of longitudinal studying across different periods in the same jurisdiction, leading to potential biased temporal trends analysis. Moreover, the age- and sex-specific prevalence of CKM syndrome were not calculated due to limited data. Secondly, substantial heterogeneity was observed in the entire analysis, even in the subgroup analysis. Combined with significantly publication bias, this study was rated as “low certainty of evidence” according to the GRADE approach. Thirdly, there is no standard classification guideline used in clinical setting, and studies about clinical indicators of risk stratification were limited, so more studies are warranted in the future for further validation.^52^ Fourthly, the inter-regional comparison might be influence by different study period, diagnosis criteria and other unknown factors. Lastly, although EAPC provides a global landscape of the average annual change in CKM syndrome prevalence across a specific timeframe, this method might be not be able to reflect tiny variation within a small-time range. Moreover, the quality of EAPC estimation depends on the quality of underlying data. Therefore, more studies on other regions are crucial to draw a full picture of global burden of CKM syndrome.^54^

## Conclusion

This study provides a comprehensive and latest analysis of the prevalence of CKM syndrome and its temporal trends. Even though a slight decreased trend was observed in this study, the relatively high pooled estimate calls for more attention to designing effective risk stratification and screening strategies. The substantial differences of prevalence of CKM syndrome across SDI levels, countries and data sources, as well as trend with sex ratio and average age imply potential risk factors of CKM syndrome, but need more supporting studies to verify. The public health policies aimed at reducing the stage 2 patients, in other words, the prevalence of MS, might be more effective in mitigating the burden of CKM syndrome. Finally, more comprehensive epidemiological studies in other regions, such as Africa and South America, as well as clinical biomarkers of risk stratification of CKM syndrome, are needed.

## Data sharing

The summary table of extracted data from the included studies is provided in **Table 1** and **Table 2**. All datasets generated and analyzed, including the search strategy, data extracted, and quality assessment, are available in the Article and on request from the corresponding author (JDZ).

## Guarantor Statement

All authors approved the final version of the manuscript. J.J. Li and J.D. Zhou are the guarantors of this work and, as such, had full access to all the data in the study and takes responsibility for the integrity of the data and the accuracy of the data analysis.

## Author Contributions

Data analysis: JJL, JDZ

Data review: JJL, LL, JDZ

Data acquisition: JJL, JDZ

Data interpretation: JJL, YZZ, QZG, TTZ, FFJ, PT, RH, WBC, JDZ

Critical revision of the manuscript: JJL, YZZ, QZG, TTZ, FFJ, KT, PT, DJY, RH, WBC, QQZ, JDZ

Supervision: LL, JDZ

Manuscript writing: JJL, JDZ

Manuscript revision: JJL, YZZ, QZG, TTZ, FFJ, PT, RH, WBC, QQZ, JDZ, KT

---

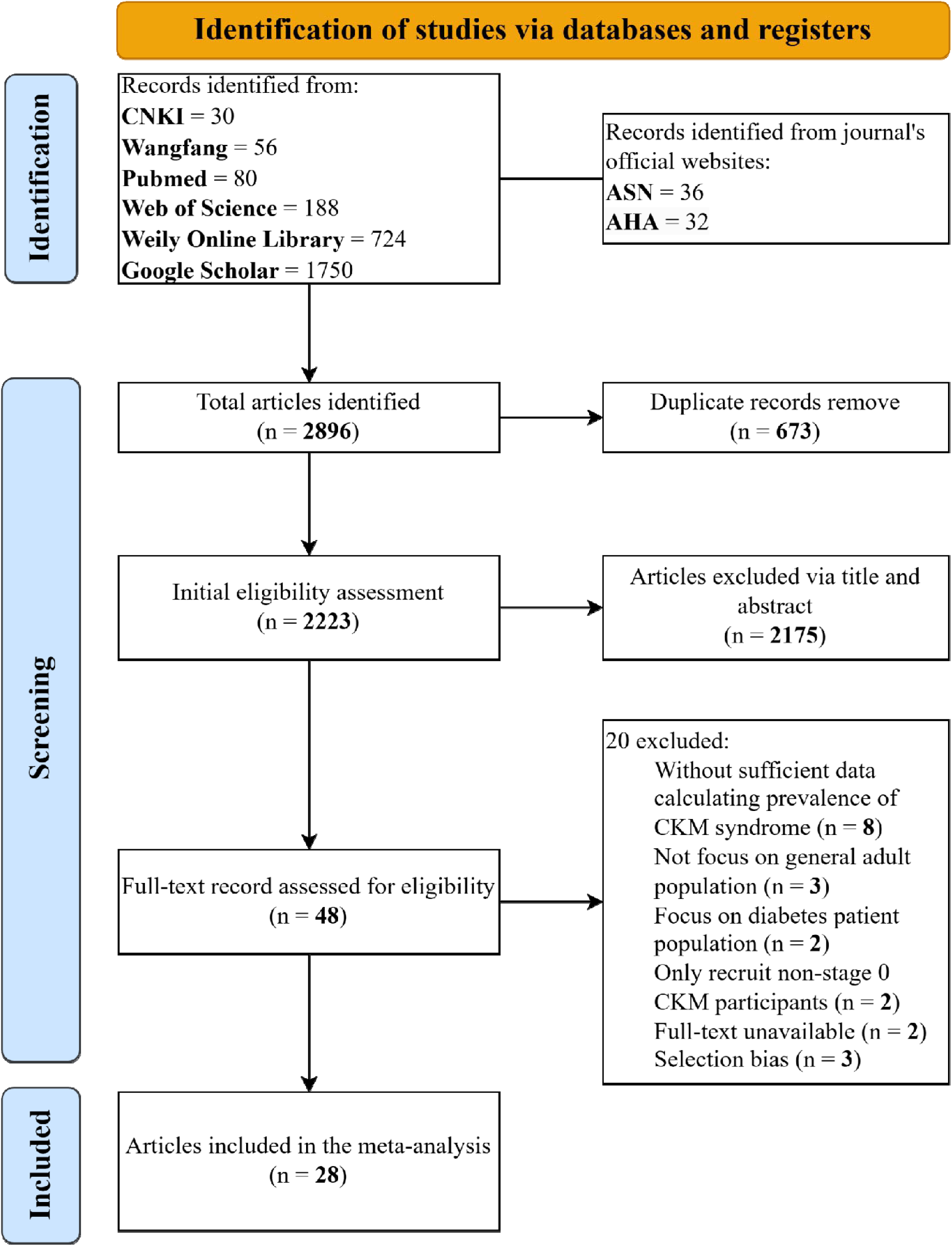

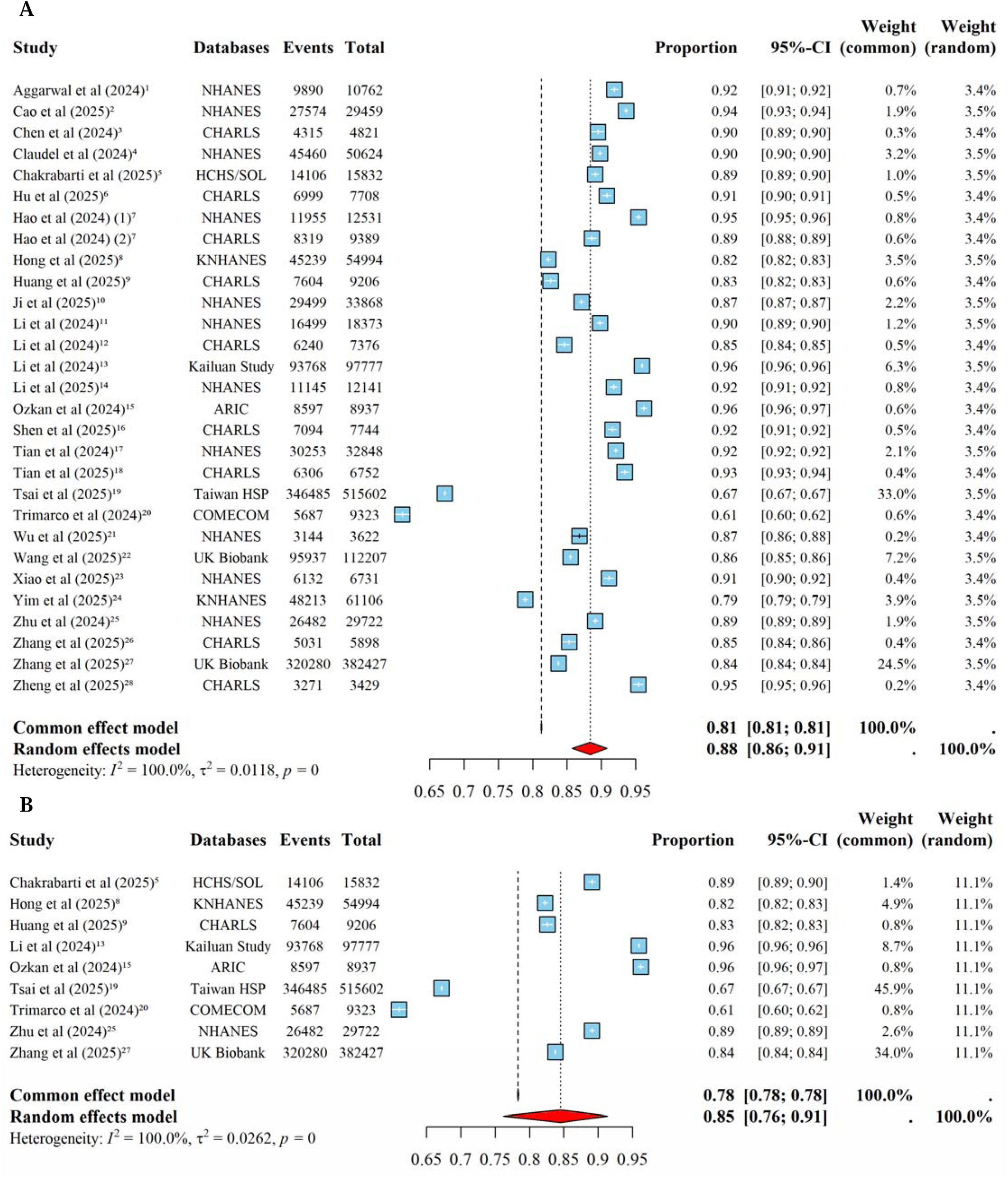

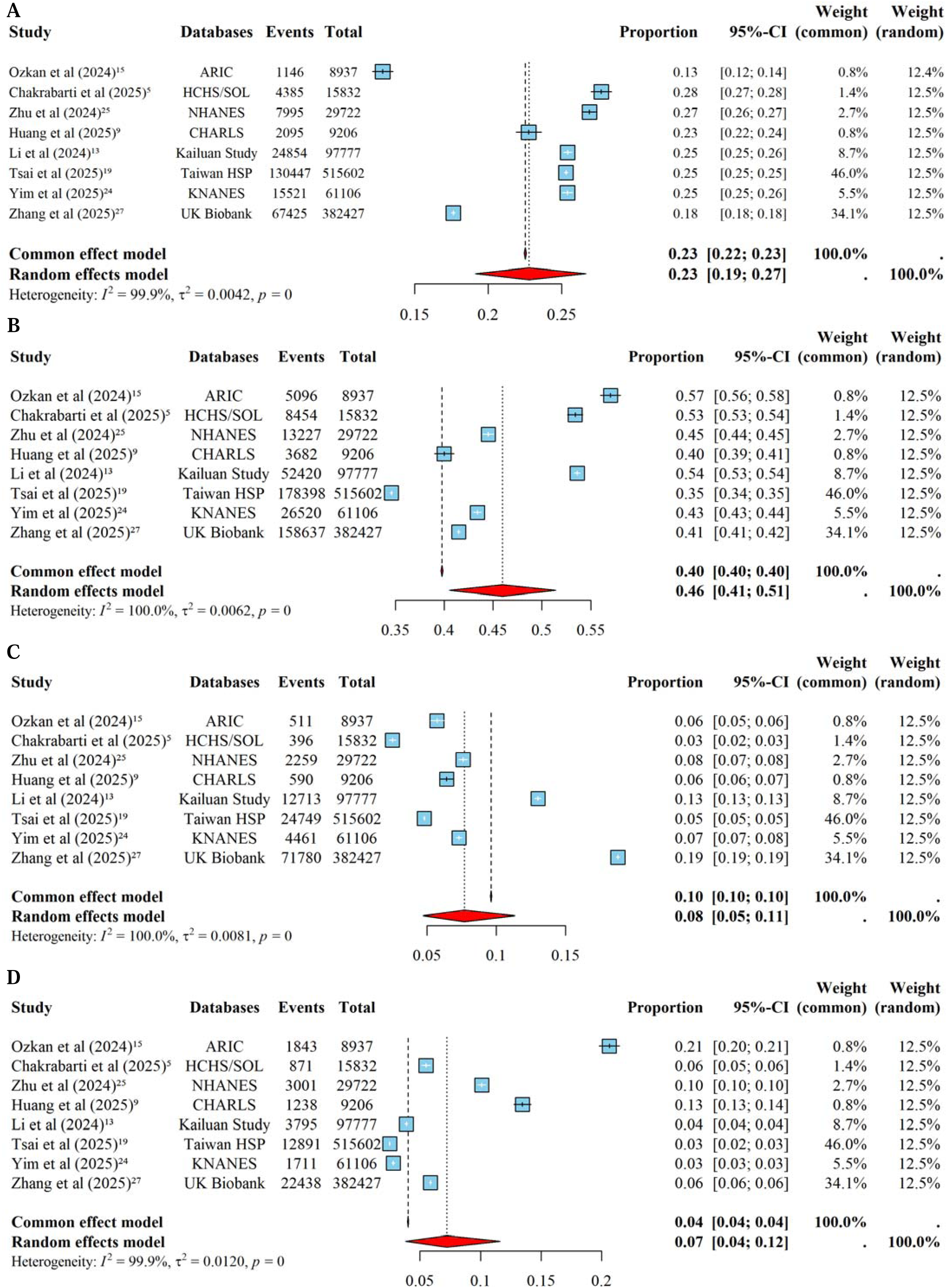

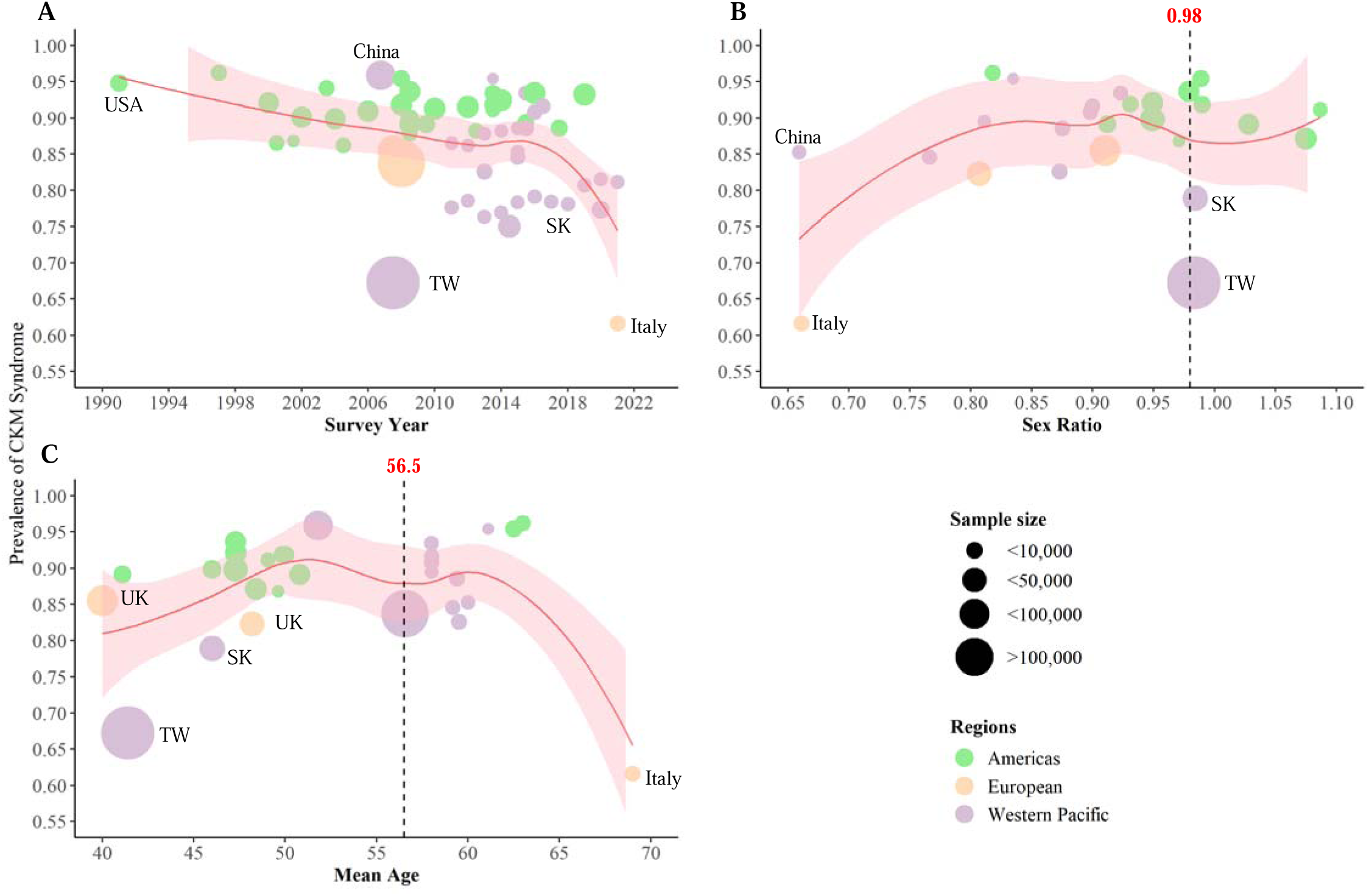

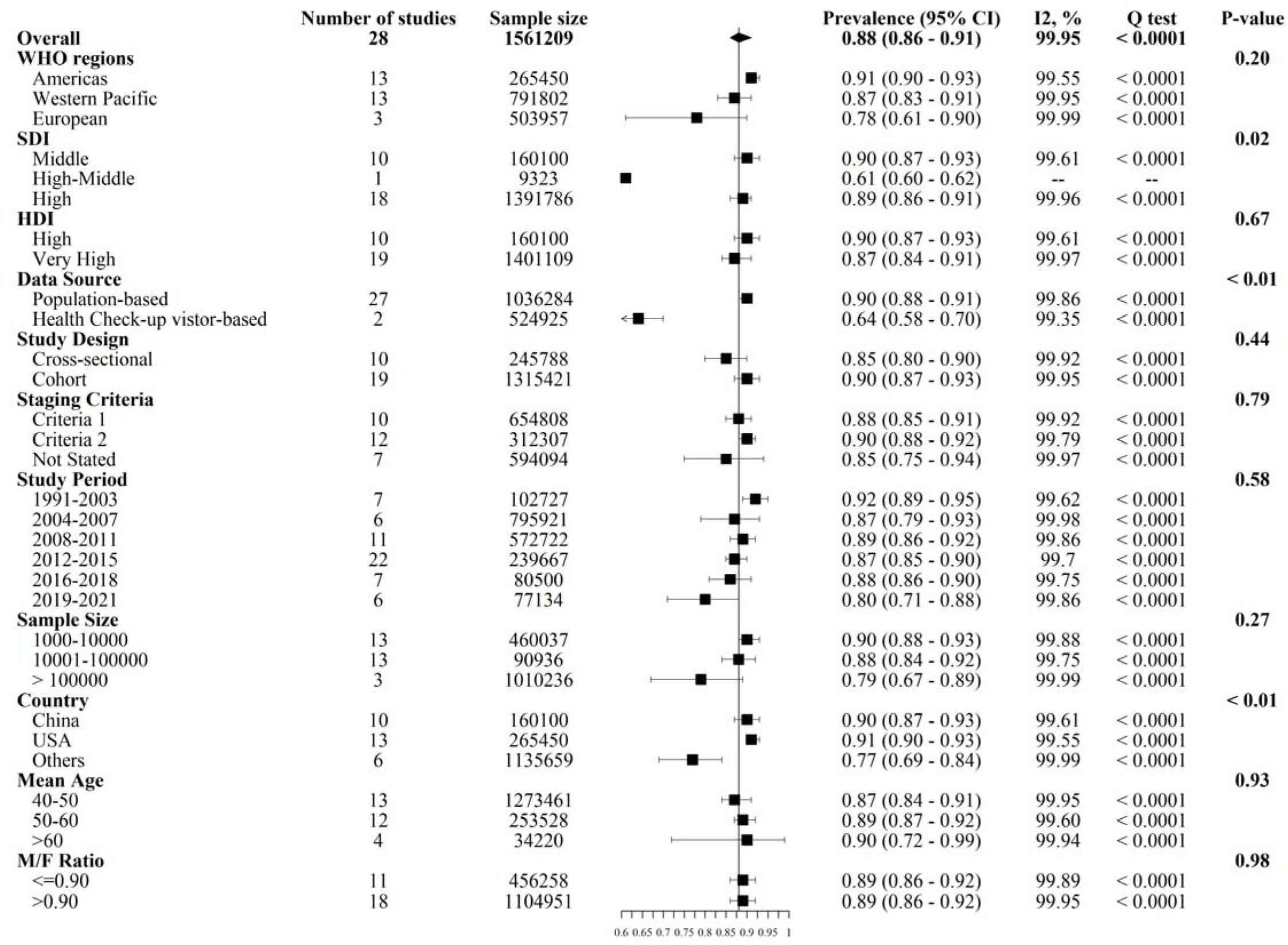

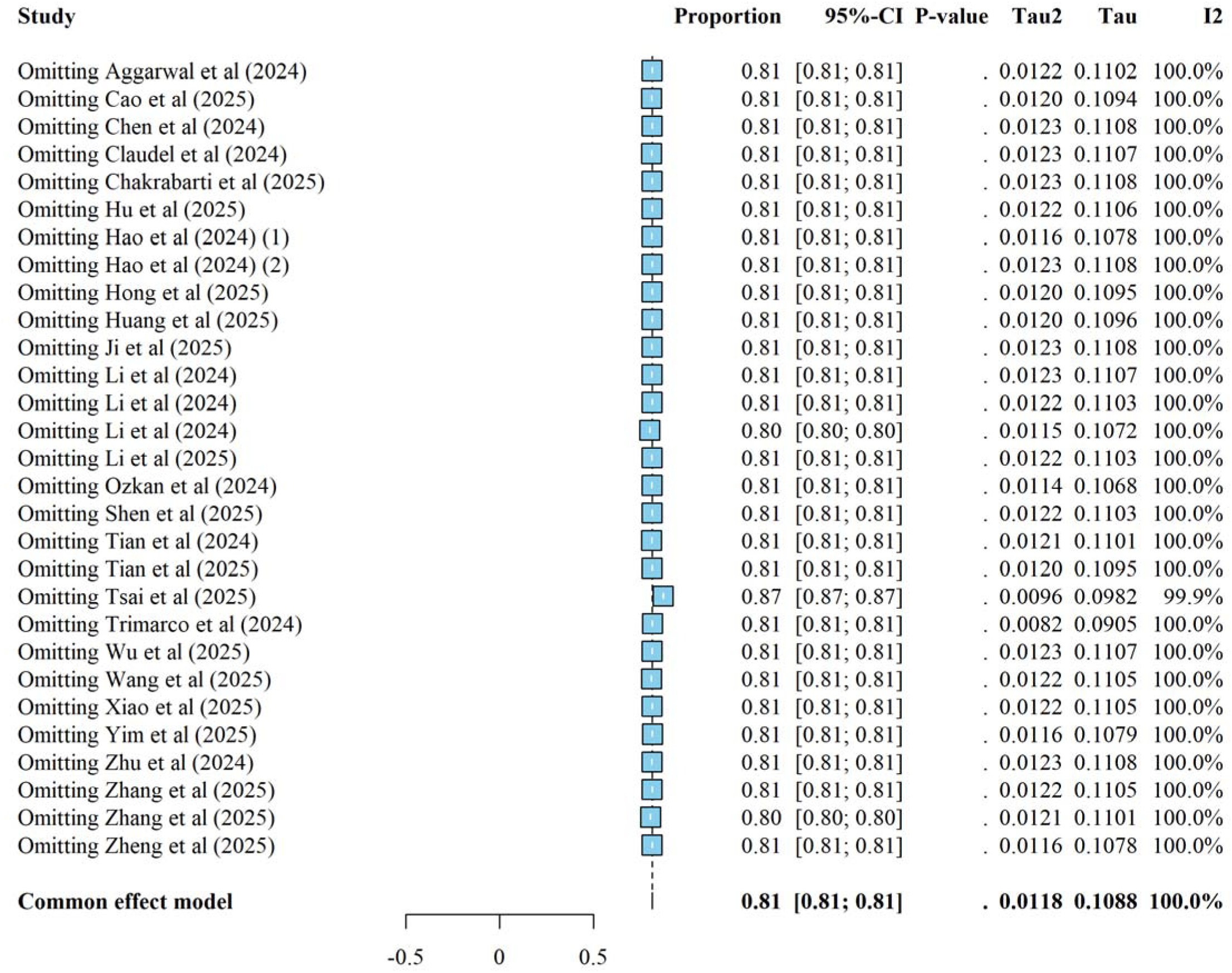

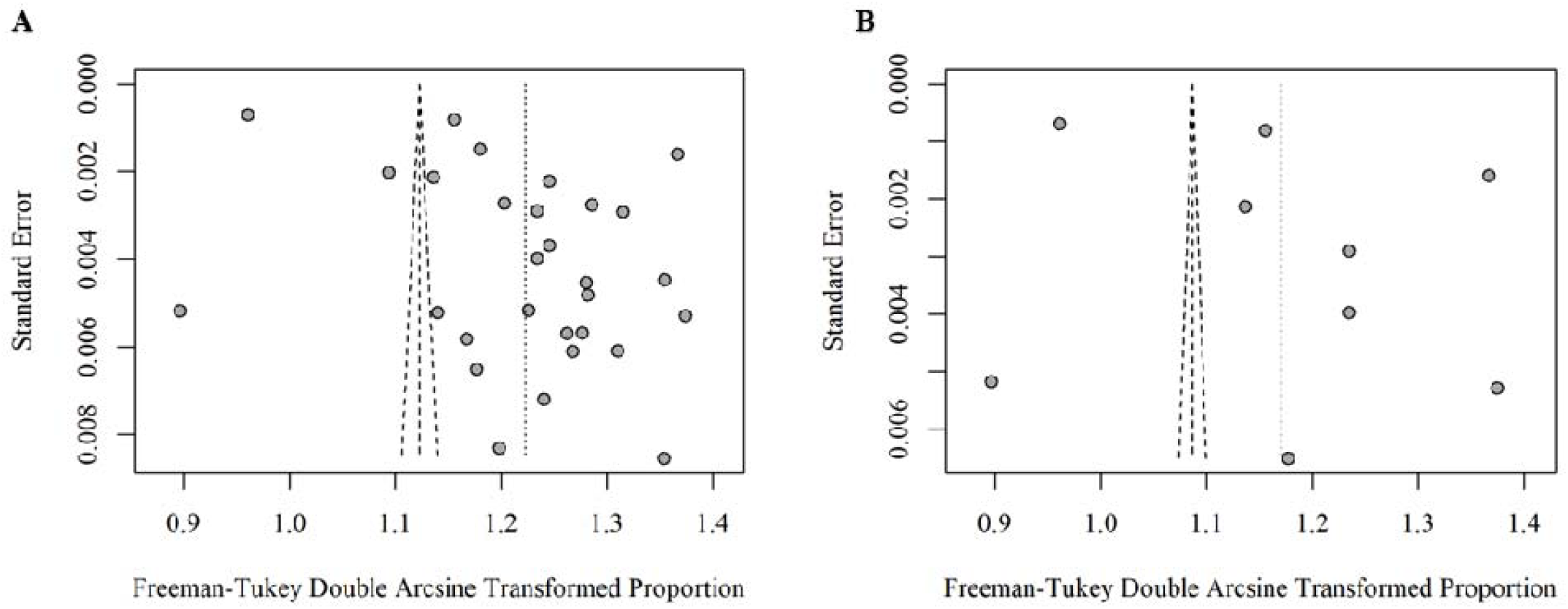

