## Appendix for "Regional Prevalence, Stage Distribution, and Temporal Trends of Cardiovascular-Kidney-Metabolic Syndrome in the Americas, Europe and Western Pacific: A Systematic Review and Meta-Analysis"

#### Context

**Table S1. Search Terms in Each Databases**

| **PubMed (1971 to April 2025)** |
|---|
| 1. “Cardiovascular Kidney Metabolic Syndrome”[Mesh]<br>2. (((((((((CKM[Title/Abstract]) OR (CKM Syndrome[Title/Abstract])) OR (New CKM syndrome[Title/Abstract])) OR (Cardiovascular-Kidney-Metabolic Syndrome[Title/Abstract])) OR (cardiovascular kidney metabolic syndrome[Title/Abstract])) OR (cardiovascular-kidney-metabolic syndrome[Title/Abstract])) OR (Cardiovascular-kidney-metabolic syndrome[Title/Abstract])) OR (CKM health[Title/Abstract])) OR (Cardiovascular Comorbidity[Title/Abstract])) OR (Metabolic Comorbidity[Title/Abstract])) OR "cardio-renal-metabolic"[Title/Abstract]))<br>3. “Cardiovascular Kidney Metabolic Syndrome”[Mesh] OR (((((((((CKM[Title/Abstract]) OR (CKM Syndrome[Title/Abstract])) OR (New CKM syndrome[Title/Abstract])) OR (Cardiovascular-Kidney-Metabolic Syndrome[Title/Abstract])) OR (cardiovascular kidney metabolic syndrome[Title/Abstract])) OR (cardiovascular-kidney-metabolic syndrome[Title/Abstract])) OR (Cardiovascular-kidney-metabolic syndrome[Title/Abstract])) OR (CKM health[Title/Abstract])) OR (Cardiovascular Comorbidity[Title/Abstract])) OR (Metabolic Comorbidity[Title/Abstract])) OR "cardio-renal-metabolic"[Title/Abstract]))<br>4. “Prevalence”[Mesh]<br>5. ((((((((Prevalences[Title/Abstract]) OR (Period Prevalence[Title/Abstract])) OR (Period Prevalences[Title/Abstract])) OR (Prevalence, Period[Title/Abstract])) OR (Point Prevalence[Title/Abstract])) OR (Point Prevalences[Title/Abstract])) OR (Prevalence, Point[Title/Abstract]) OR (Burden[Title/Abstract])) OR (Epidemiology[Title/Abstract]))<br>6. “Prevalence”[Mesh] OR (((((((((CKM[Title/Abstract]) OR (CKM Syndrome[Title/Abstract])) OR (New CKM syndrome[Title/Abstract])) OR (Cardiovascular-Kidney-Metabolic Syndrome[Title/Abstract])) OR (cardiovascular kidney metabolic syndrome[Title/Abstract])) OR (cardiovascular-kidney-metabolic syndrome[Title/Abstract])) OR (Cardiovascular-kidney-metabolic syndrome[Title/Abstract])) OR (CKM health[Title/Abstract])) OR (Cardiovascular Comorbidity[Title/Abstract])) OR (Metabolic Comorbidity[Title/Abstract])) OR "cardio-renal-metabolic"[Title/Abstract])) AND “Prevalence”[Mesh] OR ((((((((Prevalences[Title/Abstract]) OR (Period Prevalence[Title/Abstract])) OR (Period Prevalences[Title/Abstract])) OR (Prevalence, Period[Title/Abstract])) OR (Point Prevalence[Title/Abstract])) OR (Point Prevalences[Title/Abstract])) OR (Prevalence, Point[Title/Abstract]) OR (Burden[Title/Abstract])) OR (Epidemiology[Title/Abstract])) |
| **Wiley Online Library (1997 to April 2025)** |
| ("CKM" OR "CKM Syndrome" OR "New CKM syndrome" OR "Cardiovascular-Kidney-Metabolic Syndrome" OR "cardiovascular kidney metabolic syndrome" OR "cardiovascular-kidney-metabolic syndrome" OR "Cardiovascular-kidney- |

| |
|---|
| metabolic syndrome" OR "CKM health" OR "Cardiovascular Comorbidity" OR "Metabolic Comorbidity" OR "cardio-renal-metabolic") AND ("Prevalence" OR "Prevalences" OR "Period Prevalence" OR "Period Prevalences" OR "Prevalence, Period" OR "Point Prevalence" OR "Point Prevalences" OR "Prevalence, Point" OR "Burden" OR "Epidemiology") |
| **Web of Science (2004 to April 2025)** |
| "(Cardiovascular - Kidney - Metabolic Syndrome[Mesh Major Topic] OR CKM Syndrome OR New CKM syndrome OR Cardiovascular - Kidney - Metabolic Syndrome OR cardiovascular kidney metabolic syndrome OR cardiovascular - kidney - metabolic syndrome OR Cardiovascular - kidney - metabolic syndrome) AND (Epidemiology[Mesh] OR Prevalence OR Burden)" |
| **CNKI (1999 to April 2025)** |
| ("xin shen dai xie zong he zheng" OR "xin shen dai xie" OR "xin guan shen bing" OR "dai xie xing xin shen ji bing" OR "xin shen zong he zheng" OR "xin shen ji bing" OR "xin shen shuai jie" OR "xin shen zhang ai" OR "man xing shen zang bing" OR "man xing xin gong neng bu quan") AND ("huan bing lv" OR "huan bing" OR "qi jian huan bing lv" OR "dian huan bing lv" OR "liu xing bing xue" OR "fu dan" OR "fa bing lv" OR "liu xing lv" OR "liu xing te zheng" OR "liu xing qu shi" OR "wei xian yin su" OR "bing fa zheng" OR "si wang lv") |
| **WanFang (1993 to April 2025)** |
| ("xin shen dai xie zong he zheng" OR "xin shen dai xie" OR "xin guan shen bing" OR "dai xie xing xin shen ji bing" OR "xin shen zong he zheng" OR "xin shen ji bing" OR "xin shen shuai jie" OR "xin shen zhang ai" OR "man xing shen zang bing" OR "man xing xin gong neng bu quan") AND ("huan bing lv" OR "huan bing" OR "qi jian huan bing lv" OR "dian huan bing lv" OR "liu xing bing xue" OR "fu dan" OR "fa bing lv" OR "liu xing lv" OR "liu xing te zheng" OR "liu xing qu shi" OR "wei xian yin su" OR "bing fa zheng" OR "si wang lv") |
| **ASN and ASA (2023 to April 2025)** |
| ("CKM" OR "CKM Syndrome" OR "New CKM syndrome" OR "Cardiovascular-Kidney-Metabolic Syndrome" OR "cardiovascular kidney metabolic syndrome" OR "cardiovascular-kidney-metabolic syndrome" OR "Cardiovascular-kidney-metabolic syndrome" OR "CKM health" OR "Cardiovascular Comorbidity" OR "Metabolic Comorbidity" OR "cardio-renal-metabolic") AND ("Prevalence" OR "Prevalences" OR "Period Prevalence" OR "Period Prevalences" OR "Prevalence, Period" OR "Point Prevalence" OR "Point Prevalences" OR "Prevalence, Point" OR "Burden" OR "Epidemiology") |

**Table S2. Quality assessment according to the Joanna Briggs Institute Critical Appraisal Checklist for Prevalence Studies (JBIC)**

| Study | Q1 | Q2 | Q3 | Q4 | Q5 | Q6 | Q7 | Q8 | Q9 | Quality Score |
|---|---|---|---|---|---|---|---|---|---|---|
| Aggarwal (2024) | Yes | Yes | Yes | Yes | Yes | Yes | Yes | Yes | Yes | High |
| Cao (2025) | Yes | Yes | Yes | Yes | Yes | Yes | Yes | Unclear | Yes | High |
| Chen (2024) | Yes | Yes | Yes | Yes | Yes | Yes | Yes | No | Yes | Moderate |
| Claudel (2024) | Yes | Yes | Yes | Yes | Yes | Yes | Yes | Yes | Yes | High |
| Chakrabarti (2025) | Yes | Yes | Yes | No | Yes | Yes | Yes | Yes | Yes | High |
| Du (2025) | Yes | Yes | Yes | No | Yes | Yes | Yes | Yes | Yes | High |
| Hu (2025) | Yes | Yes | Yes | Yes | Yes | No | Yes | Yes | Yes | High |
| Hao (2024) (1) | Yes | Yes | Yes | Yes | Yes | No | Yes | Yes | Yes | High |
| Hao (2024) (2) | Yes | Yes | Yes | Yes | Yes | No | Yes | Yes | Yes | High |
| Hong (2025) | Yes | Yes | Yes | Yes | Yes | Yes | Yes | Yes | Yes | High |
| Huang (2024) | Yes | Yes | Yes | Yes | Yes | No | Yes | Yes | Yes | High |
| Ji (2024) | Yes | Yes | Yes | Yes | No | Yes | Yes | Yes | Yes | High |
| Li (2024) | Yes | Yes | Yes | Yes | No | Yes | Yes | Yes | Yes | High |
| Li (2024) | Yes | Yes | Yes | Yes | Yes | Unclear | Yes | Yes | Yes | High |
| Li (2024) | Yes | Yes | Yes | Yes | Yes | Yes | Yes | Yes | Yes | High |
| Li (2025) | Yes | Yes | Yes | Yes | Yes | Yes | Yes | Yes | Yes | High |
| Ozkan (2024) | Yes | Yes | Yes | Unclear | Yes | Yes | Yes | Yes | Yes | High |
| Shen (2025) | Yes | Yes | Yes | Unclear | Yes | Yes | Yes | Yes | Yes | High |
| Tian (2024) | Yes | Yes | Yes | Yes | Yes | Yes | Yes | Yes | Yes | High |
| Tian (2025) | Yes | Yes | Yes | Yes | Yes | Yes | Yes | Yes | Yes | High |
| Tsai (2025) | Yes | Unclear | Yes | No | Unclear | Yes | Unclear | Yes | Unclear | Moderate |
| Trimarco (2024) | Yes | Unclear | Yes | No | Unclear | Yes | Yes | Unclear | Unclear | Moderate |

| | | | | | | | | | | |
|---|---|---|---|---|---|---|---|---|---|---|
| Wu (2025) | Yes | Yes | Yes | Yes | Yes | Yes | Yes | Yes | Yes | High |
| Wang (2025) | Yes | Yes | Yes | Yes | Yes | Yes | Yes | Yes | Yes | High |
| Yim (2025) | Yes | Yes | Yes | Yes | Yes | Yes | Yes | Yes | Yes | High |
| Zhu (2024) | Yes | Yes | Yes | Yes | Yes | Yes | Yes | Yes | Yes | High |
| Zhang (2025) | Yes | Yes | Yes | Yes | Yes | Yes | Yes | Yes | Yes | High |
| Zhang (2025) | Yes | Yes | Yes | Yes | Yes | Yes | Yes | Yes | Yes | High |
| Zheng (2025) | Yes | Yes | Yes | Yes | Yes | Yes | Yes | Yes | Yes | High |

**Questions:**

Q1. Was the sample frame appropriate to address the target population?

Q2. Were study participants recruited in an appropriate way?

Q3. Was the sample size adequate?

Q4. Were the study subjects and setting described in detail?

Q5. Was data analysis conducted with sufficient coverage of the identified sample?

Q6. Were valid methods used for the identification of the condition?

Q7. Was the condition measured in a standard, reliable way for all participants?

Q8. Was there appropriate statistical analysis?

Q9. Was the response rate adequate, and if not, was the low response rate managed appropriately?

**Quality Rating Criteria:**

**High**: 8–9/9 "Yes" – Minimal bias, strong methodology.

**Moderate**: 5–7/9 "Yes" – Some limitations or bias.

**Low**: ≤4/9 "Yes" – Significant flaws.

**Table S3 Results of the Grading of Recommendation Assessment, Development, and Evaluation (GRADE) Assessments.**

| No. of Studies | No. of samples | Risk of Bias [a] | Inconsistency [b] | Indirectness [c] | Imprecision [d] | Publication Bias[e] | Prevalence (95% CI) | Quality |
|---|---|---|---|---|---|---|---|---|
| 28 | 1561209 | Not serious | Serious | Not serious | Not serious | Serious | 0.88 (0.86-0.91) | Low |
| CI = Confidence Interval | | | | | | | | |
| a. No included studies were rated as low quality according to the JBIC approach. | | | | | | | | |
| b. Substantial heterogeneity was observed across included studies. | | | | | | | | |
| c. All included studies were either based on well-established databases or representative population, and were conducted across multiple regions, making them inherently generalizable. | | | | | | | | |
| d. No extreme confidence intervals were observed in the study. | | | | | | | | |
| e. Funnel plot and Egger's test displayed that substantial publication bias in the combined prevalence of CKM syndrome. | | | | | | | | |

**Table S4 PRISMA Checklist**

| Section and Topic | Item # | Checklist item | Location where item is reported |
|---|---|---|---|
| **TITLE** | | | |
| Title | 1 | Identify the report as a systematic review. | P 1 |
| **ABSTRACT** | | | |
| Abstract | 2 | See the PRISMA 2020 for Abstracts checklist. | P 2 |
| **INTRODUCTION** | | | |
| Rationale | 3 | Describe the rationale for the review in the context of existing knowledge. | P 5 |
| Objectives | 4 | Provide an explicit statement of the objective(s) or question(s) the review addresses. | P 6 |
| **METHODS** | | | |
| Eligibility criteria | 5 | Specify the inclusion and exclusion criteria for the review and how studies were grouped for the syntheses. | P 7 |
| Information sources | 6 | Specify all databases, registers, websites, organisations, reference lists and other sources searched or consulted to identify studies. Specify the date when each source was last searched or consulted. | P 7 |
| Search strategy | 7 | Present the full search strategies for all databases, registers and websites, including any filters and limits used. | P 7 |
| Selection process | 8 | Specify the methods used to decide whether a study met the inclusion criteria of the review, including how many reviewers screened each record and each report retrieved, whether they worked independently, and if applicable, details of automation tools used in the process. | P 7 |
| Data collection process | 9 | Specify the methods used to collect data from reports, including how many reviewers collected data from each report, whether they worked independently, any processes for obtaining or confirming data from study investigators, and if applicable, details of automation tools used in the process. | P 8 |
| Data items | 10a | List and define all outcomes for which data were sought. Specify whether all results that were compatible with each outcome domain in each study were sought (e.g. for all measures, time points, analyses), and if not, the methods used to decide which results to collect. | P 8 |

| Section and Topic | Item # | Checklist item | Location where item is reported |
|---|---|---|---|
| | 10b | List and define all other variables for which data were sought (e.g. participant and intervention characteristics, funding sources). Describe any assumptions made about any missing or unclear information. | P 8 |
| Study risk of bias assessment | 11 | Specify the methods used to assess risk of bias in the included studies, including details of the tool(s) used, how many reviewers assessed each study and whether they worked independently, and if applicable, details of automation tools used in the process. | P 8 |
| Effect measures | 12 | Specify for each outcome the effect measure(s) (e.g. risk ratio, mean difference) used in the synthesis or presentation of results. | P 8 |
| Synthesis methods | 13a | Describe the processes used to decide which studies were eligible for each synthesis (e.g. tabulating the study intervention characteristics and comparing against the planned groups for each synthesis (item #5)). | P 8 |
| | 13b | Describe any methods required to prepare the data for presentation or synthesis, such as handling of missing summary statistics, or data conversions. | P 8 |
| | 13c | Describe any methods used to tabulate or visually display results of individual studies and syntheses. | P 8 |
| | 13d | Describe any methods used to synthesize results and provide a rationale for the choice(s). If meta-analysis was performed, describe the model(s), method(s) to identify the presence and extent of statistical heterogeneity, and software package(s) used. | P 8 |
| | 13e | Describe any methods used to explore possible causes of heterogeneity among study results (e.g. subgroup analysis, meta-regression). | P 8 |
| | 13f | Describe any sensitivity analyses conducted to assess robustness of the synthesized results. | P 8 |
| Reporting bias assessment | 14 | Describe any methods used to assess risk of bias due to missing results in a synthesis (arising from reporting biases). | P 8 |
| Certainty assessment | 15 | Describe any methods used to assess certainty (or confidence) in the body of evidence for an outcome. | P 8 |
| **RESULTS** | | | |

| Section and Topic | Item # | Checklist item | Location where item is reported |
|---|---|---|---|
| Study selection | 16a | Describe the results of the search and selection process, from the number of records identified in the search to the number of studies included in the review, ideally using a flow diagram. | P 9 |
| | 16b | Cite studies that might appear to meet the inclusion criteria, but which were excluded, and explain why they were excluded. | P 9 |
| Study characteristics | 17 | Cite each included study and present its characteristics. | P 9 |
| Risk of bias in studies | 18 | Present assessments of risk of bias for each included study. | Appendix P 3 |
| Results of individual studies | 19 | For all outcomes, present, for each study: (a) summary statistics for each group (where appropriate) and (b) an effect estimate and its precision (e.g. confidence/credible interval), ideally using structured tables or plots. | P 10 - 20 |
| Results of syntheses | 20a | For each synthesis, briefly summarise the characteristics and risk of bias among contributing studies. | P 21 |
| | 20b | Present results of all statistical syntheses conducted. If meta-analysis was done, present for each the summary estimate and its precision (e.g. confidence/credible interval) and measures of statistical heterogeneity. If comparing groups, describe the direction of the effect. | P 21 |
| | 20c | Present results of all investigations of possible causes of heterogeneity among study results. | P 21 |
| | 20d | Present results of all sensitivity analyses conducted to assess the robustness of the synthesized results. | P 21 |
| Reporting biases | 21 | Present assessments of risk of bias due to missing results (arising from reporting biases) for each synthesis assessed. | P 21 |
| Certainty of evidence | 22 | Present assessments of certainty (or confidence) in the body of evidence for each outcome assessed. | P 21 |
| **DISCUSSION** | | | |
| Discussion | 23a | Provide a general interpretation of the results in the context of other evidence. | P 22 - 23 |

| Section and Topic | Item # | Checklist item | Location where item is reported |
|---|---|---|---|
| | 23b | Discuss any limitations of the evidence included in the review. | P 23 - 24 |
| | 23c | Discuss any limitations of the review processes used. | P 23 - 24 |
| | 23d | Discuss implications of the results for practice, policy, and future research. | P 24 |
| **OTHER INFORMATION** | | | |
| Registration and protocol | 24a | Provide registration information for the review, including register name and registration number, or state that the review was not registered. | P 7 |
| | 24b | Indicate where the review protocol can be accessed, or state that a protocol was not prepared. | NA |
| | 24c | Describe and explain any amendments to information provided at registration or in the protocol. | NA |
| Support | 25 | Describe sources of financial or non-financial support for the review, and the role of the funders or sponsors in the review. | P 9 |
| Competing interests | 26 | Declare any competing interests of review authors. | P 9 |
| Availability of data, code and other materials | 27 | Report which of the following are publicly available and where they can be found: template data collection forms; data extracted from included studies; data used for all analyses; analytic code; any other materials used in the review. | P 25 |

**Text S1 Introduction to Human Development Index and Socio-demographic Index**

To investigate the correlation between CKM syndrome prevalence and the socioeconomic development, we introduced the human development index (HDI) and socio-demographic index (SDI). The Human Development Index (HDI) is a widely recognized measure developed by the United Nations Development Programmed (UNDP) to assess a country's level of human development beyond mere economic output. Introduced in 1990, it integrates three core dimensions: health, measured by life expectancy at birth; education, evaluated through mean years of schooling for adults aged 25 and older and expected years of schooling for children; and standard of living, gauged by Gross National Income (GNI) per capita adjusted for purchasing power parity (PPP). The HDI is computed as a geometric mean of these normalized indices, yielding a score between 0 and 1, where 1 indicates the highest human development. Countries are classified into four tiers based on their HDI values: "Very High Human Development" (≥ 0.800), "High Human Development" (0.700–0.799), "Medium Human Development" (0.550–0.699), and "Low Human Development" (< 0.550).[1]

The Socio-demographic Index (SDI) is a composite metric created by the Institute for Health Metrics and Evaluation (IHME) as part of the Global Burden of Disease (GBD) study to evaluate a region's socio-economic development, particularly in relation to health outcomes.[2] It combines three key indicators: income, represented by lagged distributed income per capita; education, measured by average years of schooling for individuals aged 15 and older; and fertility, assessed via the total fertility rate for women under 25, which reflects demographic transitions tied to development. The SDI is calculated as a geometric mean of these components, scaled from 0 (lowest development) to 1 (highest development), and is often used to analyze health disparities globally. For classification, the SDI is typically divided into quintiles: "Low SDI" (bottom 20%, e.g., 0–0.2), "Low-Middle SDI" (0.2–0.4), "Middle SDI" (0.4–0.6), "High-Middle SDI" (0.6–0.8), and "High SDI" (top 20%, e.g., 0.8–1.0). These categories are relative, with high SDI regions like Western Europe contrasting with low SDI areas like parts of sub-Saharan Africa.

**Text S2 Male/female ratio and average age in the included studies**

Since the majority of the included studies did not provide sex- and age- specific prevalence data for CKM syndrome, we employed the male/female ratio and the average age of the participants in each study as proxies for sex- and age- related factors. This approach represents a preliminary exploration of two crucial risk factors, sex and aging, in the burden of CKM syndrome. By analyzing these proxies, we aimed to assess whether variations in the sex and age distributions contribute to the overall burden of CKM syndrome. If so, future research can be more confidently designed to investigate the prevalence of CKM syndrome separately in males and females, as well as across different age groups, with more comprehensive and detailed data collection strategies. This method, although a compromise due to data limitations, allows us to generate valuable preliminary insights and guide subsequent in - depth investigations.

**Text S3 Staging criteria of CKM syndrome**

Studies with different staging criteria of CKM syndrome were categorized into three groups: criteria 1 (e.g. strictly follows principles proposed by the American Heart Association (AHA) and put it into the appendix);[3] criteria 2 (e.g. also strictly follow the advisory proposed by AHA but only included in the manuscript)[4] and criteria not stated (e.g. rough description or detailed descriptions are not available)[5].

**Text S4 Estimated annual percentage change (EAPC)**

EAPC was used to explore the trends of CKM syndrome. We employed linear regression to the natural logarithm of the prevalence rate (r) to get the EAPC and its 95% CI. The final equation would be: $ln(r) = \alpha + \beta x + \varepsilon$. In where, $\alpha$ is the intercept, $\beta$ is the regression coefficient on x (such as survey year), $\varepsilon$ is the error term. EAPC is then calculated as $100 \times (e^{\beta} - 1)$.[6]

### Figure S1 Sensitivity analysis with leave-one-out approach

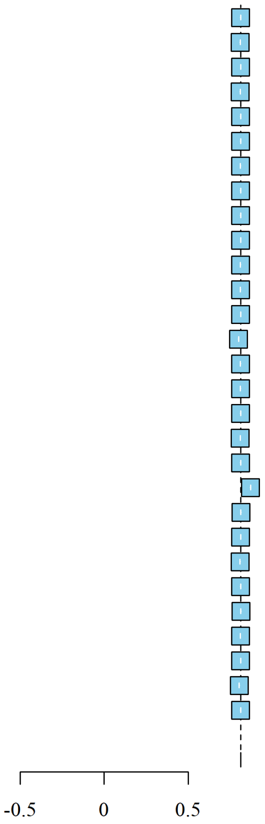

| Study | Proportion | 95%-CI | P-value | Tau2 | Tau | I2 |
|---|---|---|---|---|---|---|
| Omitting Aggarwal et al (2024) | 0.81 | [0.81; 0.81] | . | 0.0122 | 0.1102 | 100.0% |
| Omitting Cao et al (2025) | 0.81 | [0.81; 0.81] | . | 0.0120 | 0.1094 | 100.0% |
| Omitting Chen et al (2024) | 0.81 | [0.81; 0.81] | . | 0.0123 | 0.1108 | 100.0% |
| Omitting Claudel et al (2024) | 0.81 | [0.81; 0.81] | . | 0.0123 | 0.1107 | 100.0% |
| Omitting Chakrabarti et al (2025) | 0.81 | [0.81; 0.81] | . | 0.0123 | 0.1108 | 100.0% |
| Omitting Hu et al (2025) | 0.81 | [0.81; 0.81] | . | 0.0122 | 0.1106 | 100.0% |
| Omitting Hao et al (2024) (1) | 0.81 | [0.81; 0.81] | . | 0.0116 | 0.1078 | 100.0% |
| Omitting Hao et al (2024) (2) | 0.81 | [0.81; 0.81] | . | 0.0123 | 0.1108 | 100.0% |
| Omitting Hong et al (2025) | 0.81 | [0.81; 0.81] | . | 0.0120 | 0.1095 | 100.0% |
| Omitting Huang et al (2025) | 0.81 | [0.81; 0.81] | . | 0.0120 | 0.1096 | 100.0% |
| Omitting Ji et al (2025) | 0.81 | [0.81; 0.81] | . | 0.0123 | 0.1108 | 100.0% |
| Omitting Li et al (2024) | 0.81 | [0.81; 0.81] | . | 0.0123 | 0.1107 | 100.0% |
| Omitting Li et al (2024) | 0.81 | [0.81; 0.81] | . | 0.0122 | 0.1103 | 100.0% |
| Omitting Li et al (2024) | 0.80 | [0.80; 0.80] | . | 0.0115 | 0.1072 | 100.0% |
| Omitting Li et al (2025) | 0.81 | [0.81; 0.81] | . | 0.0122 | 0.1103 | 100.0% |
| Omitting Ozkan et al (2024) | 0.81 | [0.81; 0.81] | . | 0.0114 | 0.1068 | 100.0% |
| Omitting Shen et al (2025) | 0.81 | [0.81; 0.81] | . | 0.0122 | 0.1103 | 100.0% |
| Omitting Tian et al (2024) | 0.81 | [0.81; 0.81] | . | 0.0121 | 0.1101 | 100.0% |
| Omitting Tian et al (2025) | 0.81 | [0.81; 0.81] | . | 0.0120 | 0.1095 | 100.0% |
| Omitting Tsai et al (2025) | 0.87 | [0.87; 0.87] | . | 0.0096 | 0.0982 | 99.9% |
| Omitting Trimarco et al (2024) | 0.81 | [0.81; 0.81] | . | 0.0082 | 0.0905 | 100.0% |
| Omitting Wu et al (2025) | 0.81 | [0.81; 0.81] | . | 0.0123 | 0.1107 | 100.0% |
| Omitting Wang et al (2025) | 0.81 | [0.81; 0.81] | . | 0.0122 | 0.1105 | 100.0% |
| Omitting Xiao et al (2025) | 0.81 | [0.81; 0.81] | . | 0.0122 | 0.1105 | 100.0% |
| Omitting Yim et al (2025) | 0.81 | [0.81; 0.81] | . | 0.0116 | 0.1079 | 100.0% |
| Omitting Zhu et al (2024) | 0.81 | [0.81; 0.81] | . | 0.0123 | 0.1108 | 100.0% |
| Omitting Zhang et al (2025) | 0.81 | [0.81; 0.81] | . | 0.0122 | 0.1105 | 100.0% |
| Omitting Zhang et al (2025) | 0.80 | [0.80; 0.80] | . | 0.0121 | 0.1101 | 100.0% |
| Omitting Zheng et al (2025) | 0.81 | [0.81; 0.81] | . | 0.0116 | 0.1078 | 100.0% |
| **Common effect model** | **0.81** | **[0.81; 0.81]** | **.** | **0.0118** | **0.1088** | **100.0%** |

**Figure S2 Funnel plot of the all-age and sex prevalence of CKM syndrome with all included studies (A) and representative studies (B)**

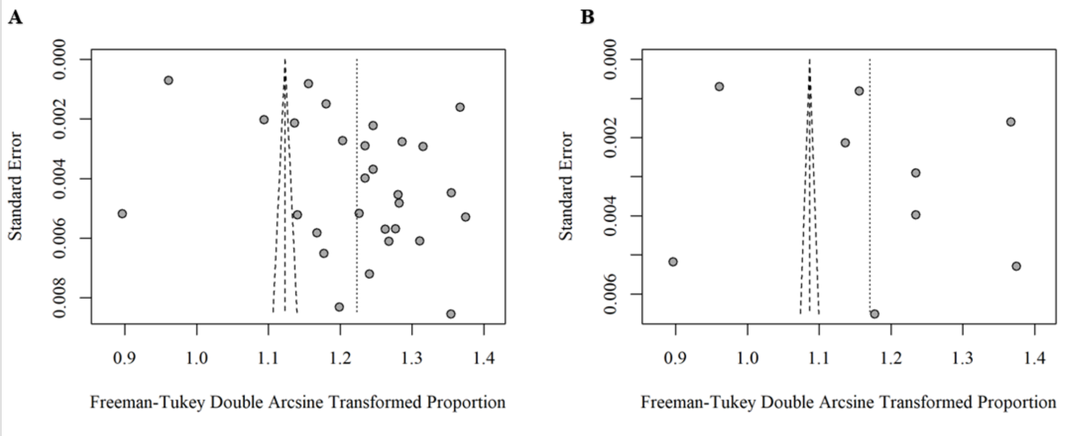
